# How Ethiopia’s elite distance runners actually train: an AI-assisted dissection of a multidimensional training structure

**DOI:** 10.64898/2026.05.29.26354013

**Authors:** Palo Galko, Abay Yisamaw, Thomas Haugen, Stephen Seiler

**Affiliations:** Independent Researcher, BambooAI Lab, Melbourne, Australia; College of Natural and Computational Sciences, Addis Ababa University, Addis Ababa, Ethiopia; School of Health Sciences, Kristiania University of Applied Sciences, Oslo, Norway; Department of Sport Science and Physical Education, University of Agder, Kristiansand, Norway

**Keywords:** human-AI collaboration, large language models, research workflow, adversarial review, citation verification, reproducibility, observational sports science, altitude training, elite distance running, Pearl’s causal hierarchy

## Abstract

**Background:** Elite East African distance runners have dominated their events for decades while training in ways that diverge sharply from Western sports-science orthodoxy. Why they are so good has never been settled with certainty. Genetics, lifelong altitude residency, and economic motivation have all been offered without a conclusive answer, partly because the day-to-day texture of the training itself has been hard to observe directly. Consumer GPS watches, now routinely used by elite athletes, leave detailed records of training that can be reconstructed at high resolution. This study takes the detailed footprint left in one cohort’s watch data and dissects it for the factors behind their performance, using generative AI to make a decomposition of this depth tractable. The pattern that finally made sense of the data came from human expertise, echoing the very way these runners are coached.

**Methods:** We followed 14 elite Ethiopian distance runners across 97 consecutive days in late 2025, drawing on 22,605 GPS segment records from their training watches together with venue and athlete information gathered in the field. The analysis was carried out through a three-phase human-AI workflow. An autonomous AI tool first searched the data across five investigator-seeded questions. A second AI system then developed selected findings into numerical claims, code, figures, and draft text under direct human guidance. A third, independent AI system stress-tested the methods, statistics, figures, and citations. Across all phases, the investigators set the direction, judged what the data could support, applied corrections and supplied the interpretation.

**Results:** The dissection reconstructs how the cohort trains. Rather than working along a single intensity axis, the athletes spread their training across venues that each combine a particular altitude, surface, and terrain, with long aerobic running on the gentler mid-altitude roads, varied trail running at the high-altitude sites where the route changes from session to session, structured repetitions on the track, and easy recovery spread across the rest. Intensity is set by effort rather than by fixed pace or heart-rate targets, so altitude, surface, and intensity move together and cannot be cleanly separated by statistics. Essentially all the key sessions are morning group runs, and within the long ones the athletes settle into fitness-matched pace bands, so the group itself does the individualisation that Western coaching delivers through prescribed individual zones. Measured against the standard half-marathon and marathon pace scale, the intensity distribution is dominated by easy running with only a small moderate share and leans polarised once altitude is accounted for, matching the training logs of world-class runners recorded at altitude. Taken alone, it looks unremarkable, and that single-axis summary is a projection that hides how the work is placed, which only the multidimensional view recovers. All of it condenses into a compact map with five axes, altitude, surface, intensity, session structure, and session duration, drawn as three lattices on a shared altitude-and-surface grid, which the coaches arrive at by experience and the cohort concentrates into a handful of combinations.

**Conclusions:** In this cohort, the GPS-watch data revealed a multidimensional training pattern in which intensity, altitude, surface, terrain, and group execution varied together rather than falling along a single intensity axis. The multidimensional framework developed to describe the data should be understood as a descriptive model of the regularities visible in that record, with no implication that the coaches used a formal scheme. Its value is that it makes part of an experiential coaching practice measurable. The coaches’ tacit knowledge shaped the training, and the training left a structured footprint in the data. The AI-assisted workflow helped expose and verify that footprint, but the organising interpretation depended on human domain expertise. The findings therefore describe how this cohort trained in this setting, and any application beyond it would require judgement grounded in the athletes, coaches, venues, and constraints of the new context.

## Background

For more than four decades, the longest events in distance running have been dominated by a small number of athletes from the East African highlands, and Ethiopia sits close to the centre of that record. The question of why has been asked directly in the literature for almost as long, and it remains open [1]. Several explanations have been offered, each with evidence behind it and none satisfying in isolation: a genetic inheritance suited to endurance, the physiological consequences of living and training at altitude from childhood, and the powerful motivation that running provides as a route out of poverty. What ties these athletes together is harder to locate than any single one of those factors, and part of the difficulty is that they train in ways that depart sharply from the orthodoxy taught in Western sports science [2, 3].

What is actually known about that training has been gathered mostly indirectly. The daily practice of East African runners has been described in the sport science literature through training diaries, interviews, and the occasional laboratory visit [1, 4, 5], and ethnographically through accounts of how the runners themselves relate to effort and to self-tracking technology [19]. It has usually been described through the categories Western sports science already had to hand: weekly volume, the balance of hard and easy days, the distribution of training across intensity zones, and the effects of altitude [6, 7]. These are useful summaries, but they are coarse and indirect, and they describe a distinctive practice through categories borrowed from elsewhere. The fine structure of the training, how individual sessions are built, where they are run, how pace and recovery are managed, and how all of this interacts with geography, training conditions, and culture, has stayed largely out of direct view.

Consumer GPS watches can now be used to log pace, heart rate, altitude, and location continuously, in the field, at a resolution that once required dedicated research equipment. For a group whose training has been seen only in fragments, this offers a near-complete record of what was actually done, day after day, across a season.

The record is also large and tangled, which is where a second instrument enters. Tens of thousands of segment-level observations, each carrying several interacting variables, are difficult to dissect by hand, and generative AI now makes a decomposition of that size tractable. It can search the data, write and run the analysis, and draft the account at a speed that opens questions which would otherwise stay closed. The AI tools can carry the labour of the dissection, but the judgement of what to look for, and of what the data is and is not saying, stays with the investigators doing the work.

This paper applies that combination to a single cohort: 14 elite Ethiopian distance runners, six men and eight women, recorded across 97 consecutive days of training in late 2025, producing 22,605 segment records, with venue and athlete information supplied from the field. The work is observational and descriptive: one cohort, one competitive season, no comparison group, and no laboratory measurement. The account that follows reconstructs, in high resolution, how this particular group trained, and the larger question of why any group comes to dominate is beyond what a design like this can settle. What it offers is a more direct look at the texture of elite Ethiopian training than the indirect methods have allowed, and an attempt to read, in the trace the athletes left behind, the shape of the practice that produced it.

## Methods

This study used generative AI as a working instrument inside an empirical investigation. This section first describes how the work was divided between the AI tools and the investigators, and then sets out the cohort, the data, and the analytical methods themselves. To locate the division precisely, we borrow a simple vocabulary for the kinds of thinking an investigation requires.

### A vocabulary for dividing the work

The computer scientist Judea Pearl distinguishes three kinds of cognitive work, ranking them by the questions each can answer [8]. The first is **association**: which patterns occur together. Given training data, the hardest sessions may fall on the same days each week, or one surface may show a higher heart rate at a given pace than another. Current language models are strong at this kind of work, drawing on patterns learned from large bodies of text and code to search a dataset, write and run analysis, draft prose, and match a question to relevant literature. The second is **intervention**: what changes if something is altered. In an investigation this means choosing which line of enquiry to drop, demanding a sensitivity check, changing a specification when a result looks clean for the wrong reason, and holding a long study together across many sessions. The third is **counterfactual reasoning**: what would hold under conditions the data does not directly show, including whether a finding generalises and whether a framework should apply where the data cannot test it. This is the work of interpretation and judgement. The three name kinds of work rather than stages, and a single working session typically contains all of them.

Across these, current large language models are strong at the first kind of work, can assist with the second under direct guidance, and leave the third with the investigator. A language model can draw only on the concepts and relations present in its training data. It can recombine what is already there to expose new patterns, but it does not, on its own, reach for an organising framework built on terms its training did not contain [9]. When a dataset needs to be reorganised around axes the model has never seen, that reorganisation comes from outside the model. Supplying such a framework is the highest part of the human contribution, and it sits just above the top rung.

### The three-phase workflow

The investigation divided this work across three phases, each with its own input, output, and means of catching error. The first phase used Delv-e, an autonomous data-exploration tool [10], run five times against the cleaned dataset from five seeded questions. Within a fixed iteration budget it generated and tested candidate findings, wrote and ran its own code, and adapted across iterations without intervention, returning a structured briefing of promising directions, foreclosed paths, and open questions. This is associative search at scale. The second phase worked through selected candidates one claim at a time under direct human guidance, using Claude Opus 4.7 [11]. Each finding was stated, tested in code against the data, inspected, compared against the literature, and then accepted, narrowed, or set aside. The model supplied code, draft prose, and recognition of statistical structure; the investigator supplied the steering and the judgement of what each result meant, and kept the investigation coherent from session to session. The third phase passed the drafted work to an independent model, GPT-5.4 [12], with no part in producing it, to search for statistical failures, mismatches between prose and figures, citation errors, and overclaims, on the principle that a system is least sensitive to its own familiar mistakes. The investigator ran as a continuous channel across all three phases, holding the top of the ladder throughout. Figure 1 maps the phases against the kinds of work, and marks the point where the AI tools reach their limit and the investigator takes over.

**Figure 1.**
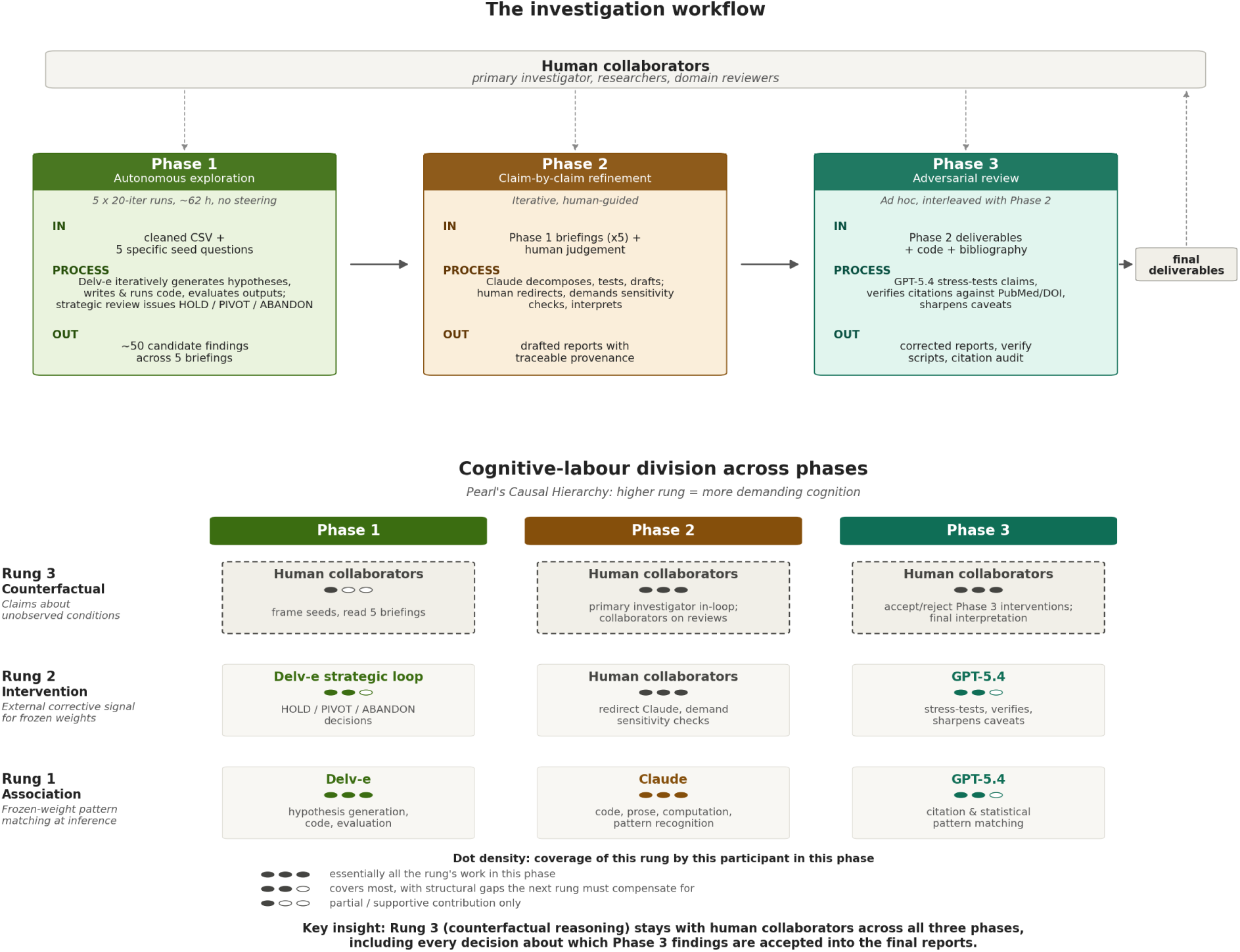
The three-phase workflow (top) and how the work divides across Pearl’s three kinds of cognitive work (bottom). The associative, lower-rung work is carried increasingly by the AI tools; the top rung, including every decision about what the data can support and the supplying of any new organising framework, stays with the investigators across all three phases.

### Cohort and data

The cohort is 14 elite Ethiopian distance runners, six men and eight women, spanning McKay Tier 4 and Tier 5 [13], recorded across 97 consecutive days between September and December 2025. Most are marathon or half-marathon specialists, with one 10 km road specialist. The window overlapped the competitive season for most of them, with races in Amsterdam, Berlin, Kolkata, Toronto, Abu Dhabi, and Košice. Their Coros watches produced 22,605 segment records carrying pace, heart rate, altitude, location, and elapsed time. Three sources were combined: the GPS segment files, athlete metadata and personal-best times, and a reference list of eight training venues spanning 2,065 to 3,100 m across six surfaces, with field context supplied by a sports scientist working in Ethiopia. Table 1 lists each athlete with sex, event, tier, and the summary measures used below. Personal-best times are withheld to protect anonymity.

**Table 1.** The 14 athletes, with sex, primary event, McKay performance tier, age range, body mass index, percentage gap to the world record in their event (%WR), and the number of recorded training segments at home in Ethiopia and abroad. Ages are given in five-year ranges and the world-record gap in three-point bins, so that personal-best times cannot be reconstructed from the table. Athletes 03, 08, and 13 did not train abroad during the recording window.

| Athlete | Sex | Event | Tier | Age | BMI | %WR | Segments (Eth) | Segments (abroad) |
| --- | --- | --- | --- | --- | --- | --- | --- | --- |
| 01 | F | 10 km road | 4 | 21–25 | 17.2 | 3–5 | 744 | 165 |
| 02 | F | Marathon/HM | 5 | 26–30 | 18.9 | 3–5 | 945 | 132 |
| 03 | F | Marathon/HM | 5 | 26–30 | 17.9 | 3–5 | 1,233 | 0 |
| 04 | F | Marathon | 4 | 26–30 | 19.6 | 9–11 | 1,095 | 60 |
| 05 | F | Marathon | 4 | 21–25 | 17.9 | 3–5 | 1,059 | 92 |
| 06 | M | Marathon | 5 | 26–30 | 20.6 | <3 | 1,999 | 100 |
| 07 | M | Marathon | 4 | 26–30 | 18.4 | 3–5 | 1,362 | 21 |
| 08 | M | Marathon | 5 | 31–35 | 19.0 | 3–5 | 2,095 | 0 |
| 09 | M | Marathon | 4 | 21–25 | 20.2 | 6–8 | 1,229 | 70 |
| 10 | M | Marathon | 5 | 26–30 | 18.0 | <3 | 1,565 | 64 |
| 11 | F | Marathon | 4 | 26–30 | 19.0 | 6–8 | 1,112 | 144 |
| 12 | F | Marathon | 4 | 16–20 | 19.5 | 9–11 | 1,186 | 54 |
| 13 | M | Marathon | 5 | 31–35 | 19.6 | 3–5 | 1,305 | 0 |
| 14 | F | Marathon/HM | 4 | 21–25 | 19.0 | 9–11 | 1,045 | 34 |

Venues were assigned from each session’s GPS start point within a conservative radius, and left unlabelled rather than forced when no venue qualified. Surface was mapped from venue. Sessions from one athlete within two hours of each other were merged into a single bout to avoid counting split recordings as separate sessions. Laps recorded at a pace slower than 10 min/km or a distance under 500 m, reflecting walking or a paused watch rather than running, were excluded from the intensity, effort, and duration measures.

### Correcting pace for altitude and surface

The headline altitude estimate uses an iso-heart-rate strategy, comparing each athlete with themselves across environments at matched effort, which avoids the venue-allocation confound that defeats a pooled regression and absorbs two distortions in the raw data, namely that most lower-altitude running fell in race weeks and that temperature differed between settings. Surface pace costs were estimated against asphalt at matched effort and gradient.

### Terrain and courses

Route overlap was measured as the fraction of shared ground between repeat sessions of the same kind at the same venue.

### Training volume and weekly structure

Weekly volume was computed per athlete, and the cohort median was recomputed with taper and recovery weeks excluded as a robustness check.

### Intensity distribution

Intensity distribution was summarised as the share of training time spent at low, moderate, and high intensity, the standard approach in the endurance-training literature [7, 14]. There the zone boundaries are anchored to physiological thresholds from blood-lactate testing. The data in this study come from training watches, which record pace and heart rate but no lactate, so threshold-anchored zones could not be constructed. Two demarcation schemes were used instead, each built from the recorded signals. The first anchored the zones to running pace, following the three-zone scale for world-class distance runners set out by Haugen and colleagues [6], in which easy running is slower than marathon pace, moderate running falls between marathon pace at its slow edge and half-marathon pace at its fast edge, and hard running is faster than half-marathon pace. Each athlete’s marathon and half-marathon paces were taken from their personal best, and where the personal best was over a different distance the missing pace was estimated with the Riegel endurance formula [18]. The two race paces differ by only about 4%, consistent with the same authors’ observation that half-marathon and marathon speed are very close together in this population. Both paces were then adjusted to altitude by adding the iso-heart-rate correction, giving a local half-marathon and local marathon pace at each elevation. Segments faster than the local half-marathon pace counted as hard, segments between the local half-marathon and local marathon pace as moderate, and the rest as easy, each weighted by its duration.

The scale’s reference paces are defined for sea level on flat ground, which is why the altitude correction is applied, and the same scale underlies the published training logs of world-class runners at altitude that the Discussion compares against. The second anchored the zones to heart rate, after Seiler [16], taking each athlete’s maximum as the 99th percentile of their recorded values and placing the boundaries at 82% and 87% of it. Because the pace-based zones shift with the altitude correction, and that correction is uncertain, the pace scheme was computed under four corrections, 0, +0.10, +0.20, and +0.35 min/km per 1,000 m, spanning the range from no correction through the headline estimate to the upper bound of its confidence interval. With the heart-rate scheme, this gave five demarcations in all. For each, the share of time in the three zones was found, and the polarisation index [17] was computed to classify the distribution as pyramidal or polarised.

### The multidimensional lattices

For the combined picture, sessions were categorised on three axes, an intensity tier, an altitude band, and a surface, each at three levels. The intensity tier divides the running by effort at two boundaries, marathon pace and a pace-to-marathon-pace ratio of 1.30, the latter marking the trough between the two modes of the zone-free pace distribution (Figure 10), giving an easy base slower than the trough, the steady endurance running between the trough and marathon pace, and the quality running faster than marathon pace that combines the moderate and hard work of the formal three-zone scale. These were cross-tabulated to give the occupancy and the concentration of training time across the available combinations, and each available cell was given a median effort, the time-weighted median of its segments’ pace after adjustment to altitude and normalisation to the cohort marathon pace, which is shown as the cell colour, and as the smaller figure, in Figure 13. The same sessions were also arranged on the altitude-and-surface grid by two whole-session properties. The first is session structure, judged from a wider set of laps that keeps genuine walking and jogging recoveries but drops only paused-watch fragments, since a single stopped lap would otherwise inflate a session’s pace spread. Structure was read on two axes, the magnitude of the pace variation, given by its coefficient of variation, and its pattern, given by the sign of the lag-1 autocorrelation of the kilometre paces. Running with a coefficient of variation below 0.10 is continuous. The rest is interval-like rep and fartlek work when the pattern alternates, consecutive kilometres moving against each other and the autocorrelation falling at or below zero, and varied when the pattern is smooth, as under a climb or a progression, consecutive kilometres moving together and the autocorrelation staying positive. At one-kilometre lap resolution a count of pace reversals sits at chance on every surface, so the autocorrelation, which is robust to that lap-to-lap noise, carries the pattern. Because the laps are one-kilometre automatic splits, sub-kilometre terrain averages out, so continuous running carries the same low variability on soft ground as on road. The second is session duration, grouped as short below 45 minutes, medium from 45 to 75, and long above 75. Each cell of these two grids was coloured by the median of its property. The three lattices can also be resolved in time. Each cell’s running time is grouped by week, so the cell holds, for each week, the time in its bins as a stacked bar, scaled per athlete active that week, and the median of its property. Collapsing a cell over the weeks returns its whole-window value. Sessions left unlabelled at the venue step were recovered for this cross-tabulation where their recorded altitude matched a known venue, a step we term altitude-proximity augmentation, with assignments made this way flagged where they bear on a result.

### Reproducibility

All retained numerical claims were re-derived from the raw segment-level data by independent verification scripts, and the observational components follow the STROBE reporting guideline.

## Results

### Correcting pace for altitude and surface

The same effort produces a different pace depending on where the running happens, so two corrections are needed before venues can be compared.

The first is altitude. Comparing each athlete with themselves at matched heart rate, across their high-altitude home and their lower-altitude race trips, every additional 1,000 m of altitude slowed them by about 0.10 min/km at equal effort, roughly six seconds per kilometre, about 2.5% at a 4:00 min/km pace (Figure 2). The estimate rests on 19 athlete-and-reference-heart-rate comparisons across the 11 athletes who trained both at altitude and abroad, 16 of them positive, with a 95% bootstrap interval of about +0.05 to +0.35 min/km per 1,000 m from 5,000 athlete-clustered resamples. A naive between-venue regression gives roughly twenty times as much, +2.01 min/km per 1,000 m, because altitude and surface are almost perfectly confounded across the venues, with a variance inflation factor of 15.9 for altitude.

**Figure 2.**
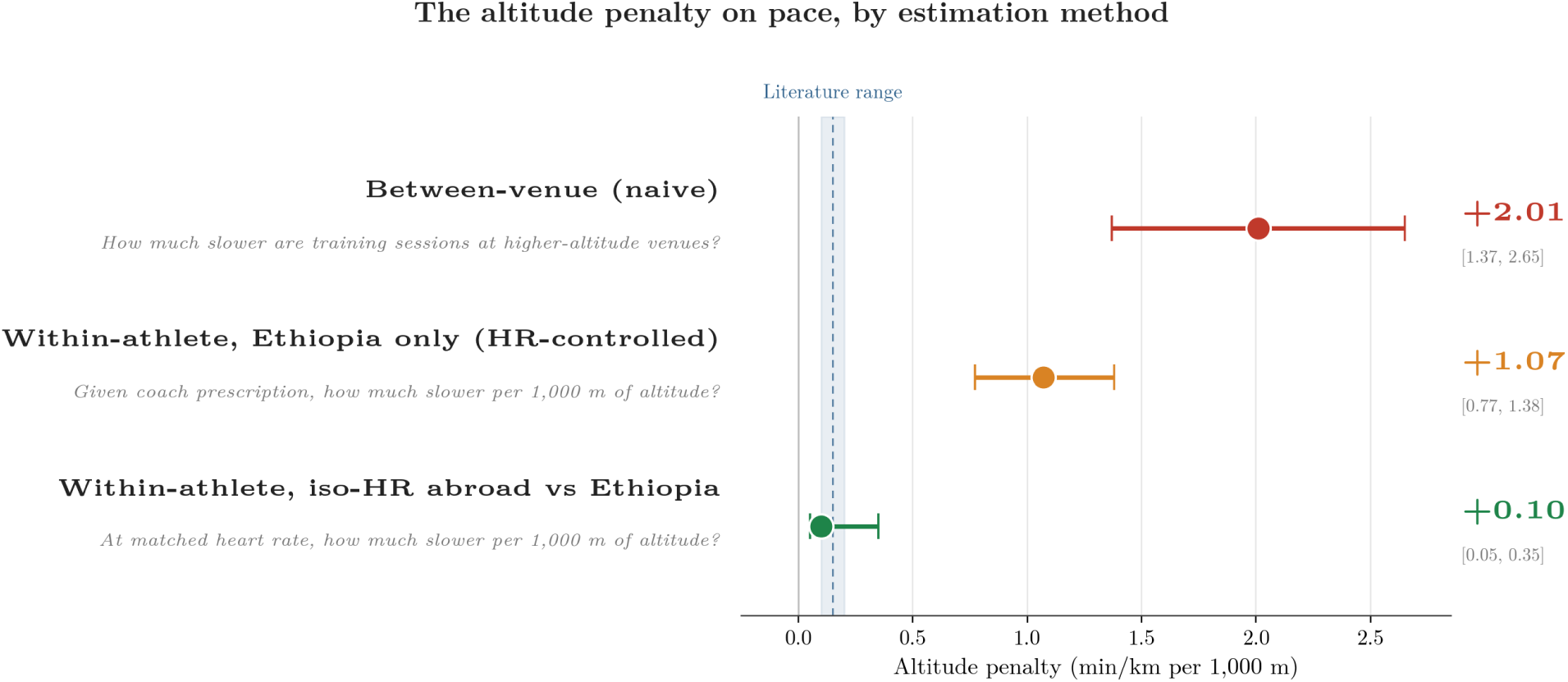
The altitude cost to pace, estimated several ways. The matched-effort estimate used in the text, about +0.10 min/km per 1,000 m, is the most conservative and the one least distorted by where and how the cohort trains. The naive between-venue estimate is about twenty times larger; the matched-effort value sits within the range published for altitude-adapted endurance runners.

The second is surface. Soft or uneven ground costs pace even at level effort and altitude. Against smooth asphalt, the dirt trails cost about 0.84 min/km, the mixed track-and-gravel about 0.48, and grass about 0.27, while the synthetic track and the firmer gravel roads ran within measurement error of asphalt (Figure 3). Each surface here belongs to a single venue. These costs come from a segment-level regression over 13,564 segments in 1,250 sessions that holds gradient constant, each additional percentage point of gradient costing about +0.24 min/km. The dirt-trail, mixed track-and-gravel, and grass costs each have intervals clear of zero, whereas the synthetic track (+0.07) and firm gravel road (−0.11) do not.

**Figure 3.**
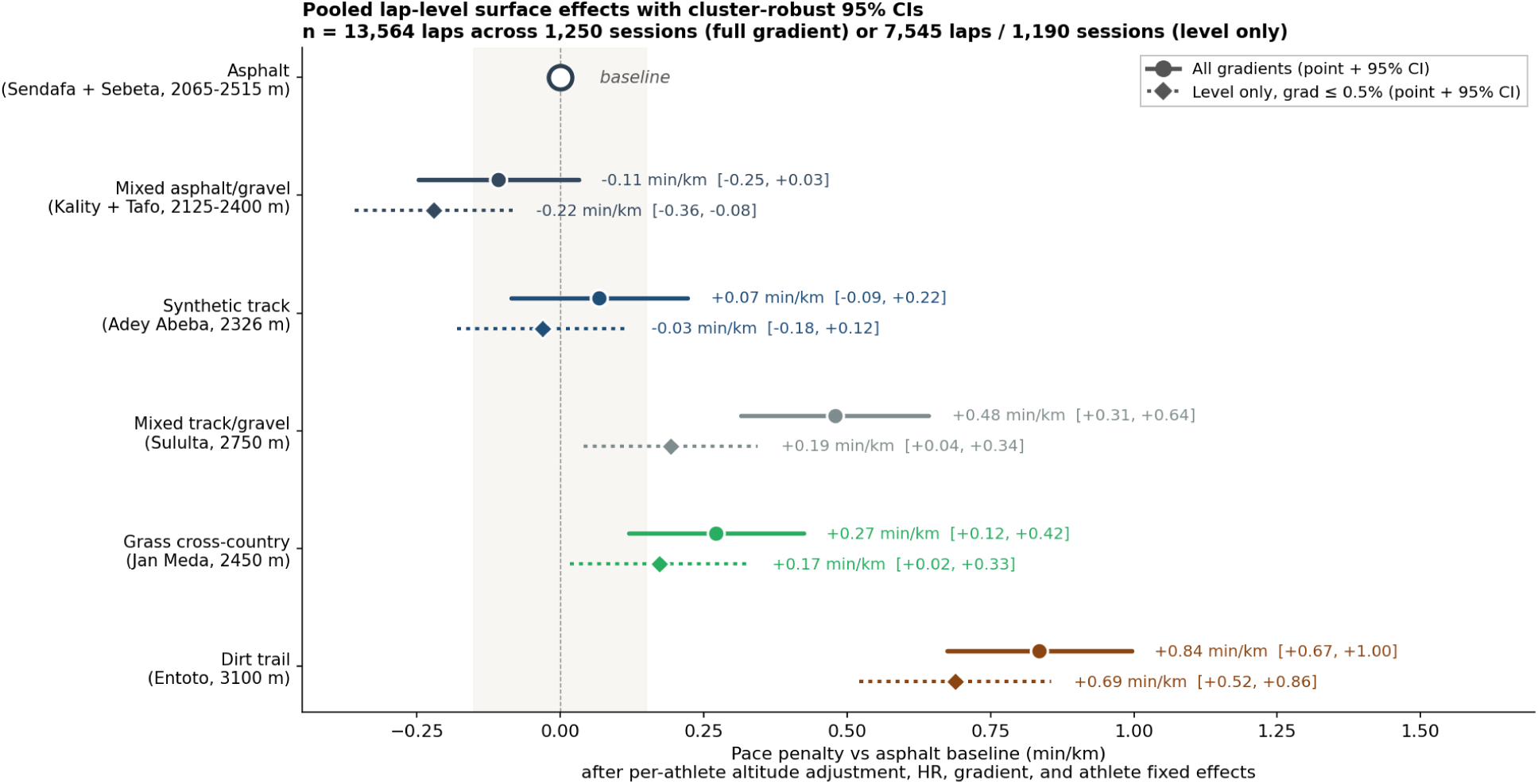
Pace cost of each surface relative to asphalt, at matched effort and gradient. Dirt trail is slowest, at about +0.84 min/km; synthetic track and firm gravel road sit within measurement error of asphalt. Each surface belongs to a single venue, so each figure reflects the place as a whole.

### The venue system and how training is shared across it

The eight venues span a wide range of height and six surfaces, and each carries its own combination of altitude, ground, and terrain. Average heart rate alone separates them, from about 156 beats per minute at the lowest, fastest road venue down to about 126 at the highest trail. Each venue is used for different kinds of work (Figure 4). The eight venues split two low, four medium, and two high by altitude, and three soft, one track, and four road by surface.

**Figure 4.**
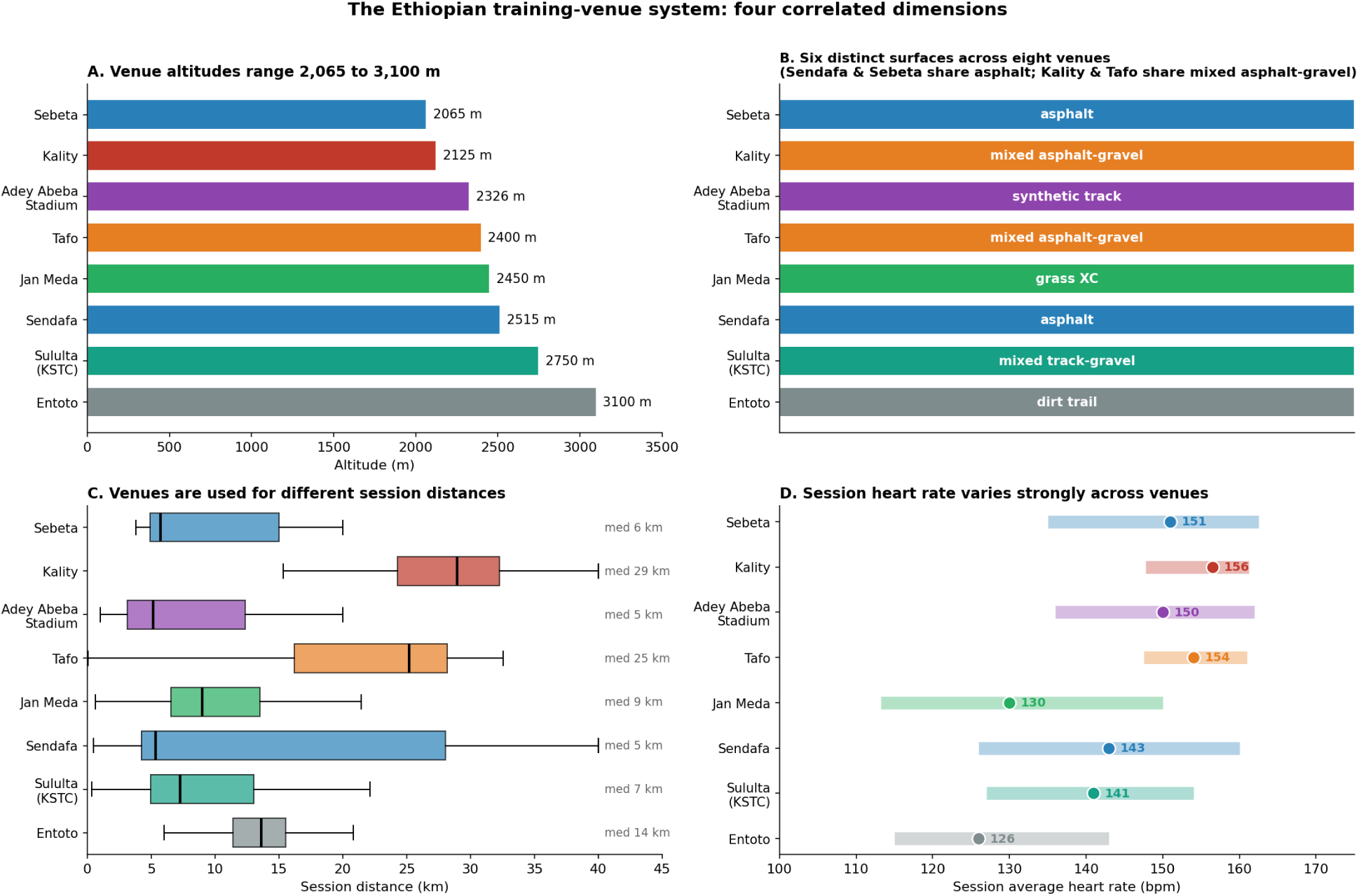
The eight training venues, set between about 2,065 and 3,100 m across six surfaces. Altitude, surface, and terrain arrive together at each venue, which is why no single one of them can be interpreted in isolation, and why a pooled regression against altitude alone overstates the altitude effect.

Training is shared across the venues in a clear pattern. The long aerobic runs concentrate on the mid-altitude gravel roads, which carry continuous running over twenty-five kilometres and more and hold about two thirds of all long-run time. The highest trail venues host the varied, rolling trail work, and the synthetic track the structured repetition sessions. The easy recovery running spreads across the remaining venues, with the choice influenced by individual proximity, and barely touches the roads, at under one percent of easy-run time.

### Terrain and courses

Knowing which venue a session ran at does not say whether the athletes cover the same ground each time. Measuring how much of the route two sessions of the same kind at the same venue share, three behaviours appear (Figure 5). On the track and the small loops the venue is the route, with nearly nine tenths of the ground shared from one session to the next. On the roads the athletes run the same path at different lengths, sharing a little under half. On the trails they cover genuinely different ground each time, sharing only a fifth to a third. Across the cohort this comes to roughly two thirds fixed-path running and one third free trail running. One venue label covers two distinct places, a central oval and a surrounding trail network used in quite different ways.

**Figure 5.**
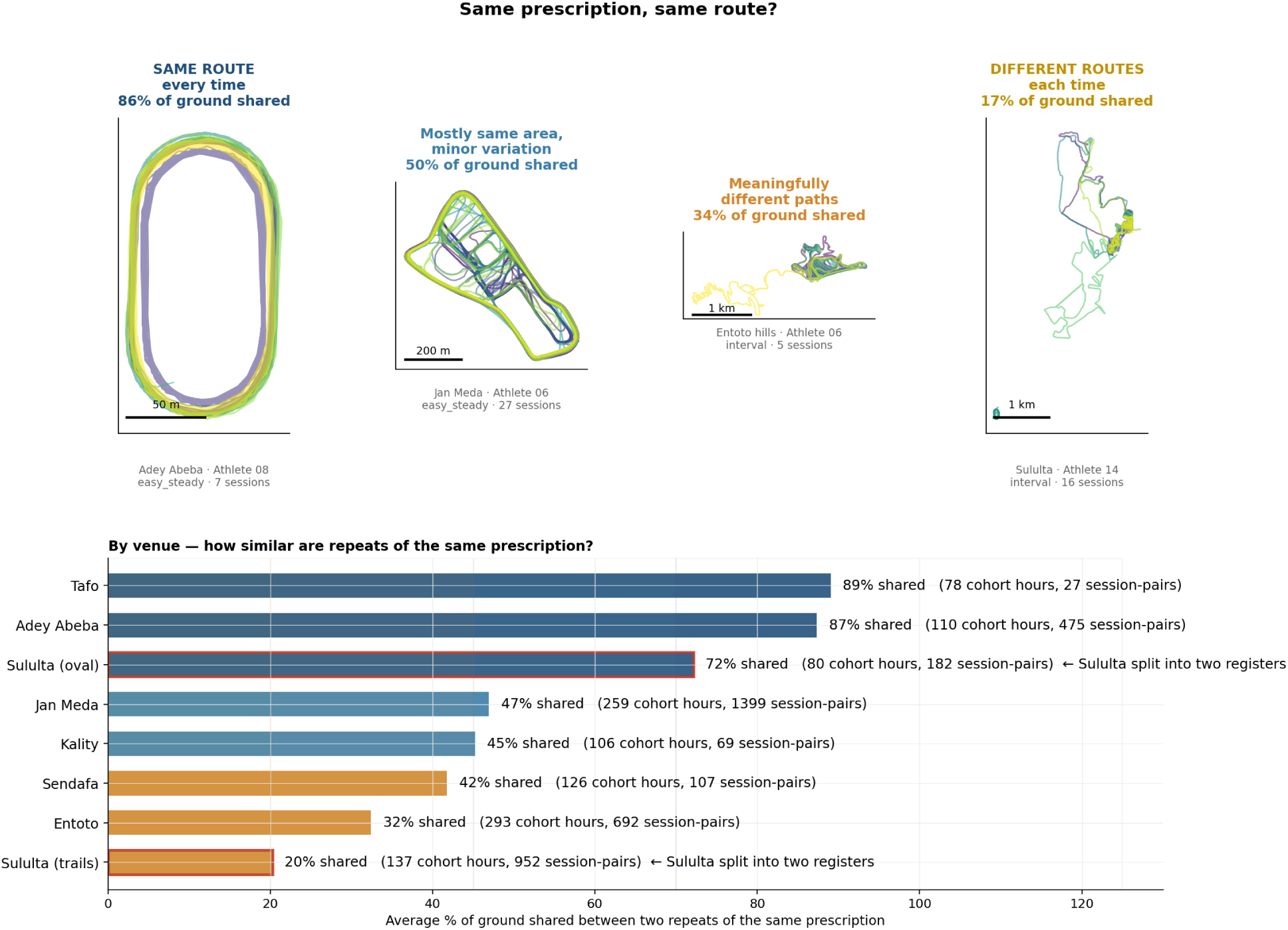
How much ground repeat sessions of the same kind share at each venue. Tracks and loops are run almost identically; roads vary in length along a fixed path; trails cover different ground each time. Across the cohort this is roughly two thirds fixed-path running and one third free trail running. The Sululta label is shown split, having concealed two distinct areas.

The terrain at each venue differs (Figure 6). The tracks are flat. The roads carrying the long runs are gently rolling. The high trails climb almost continuously or undulate throughout. The average heart rate at the highest trail is the lowest of any venue.

**Figure 6.**
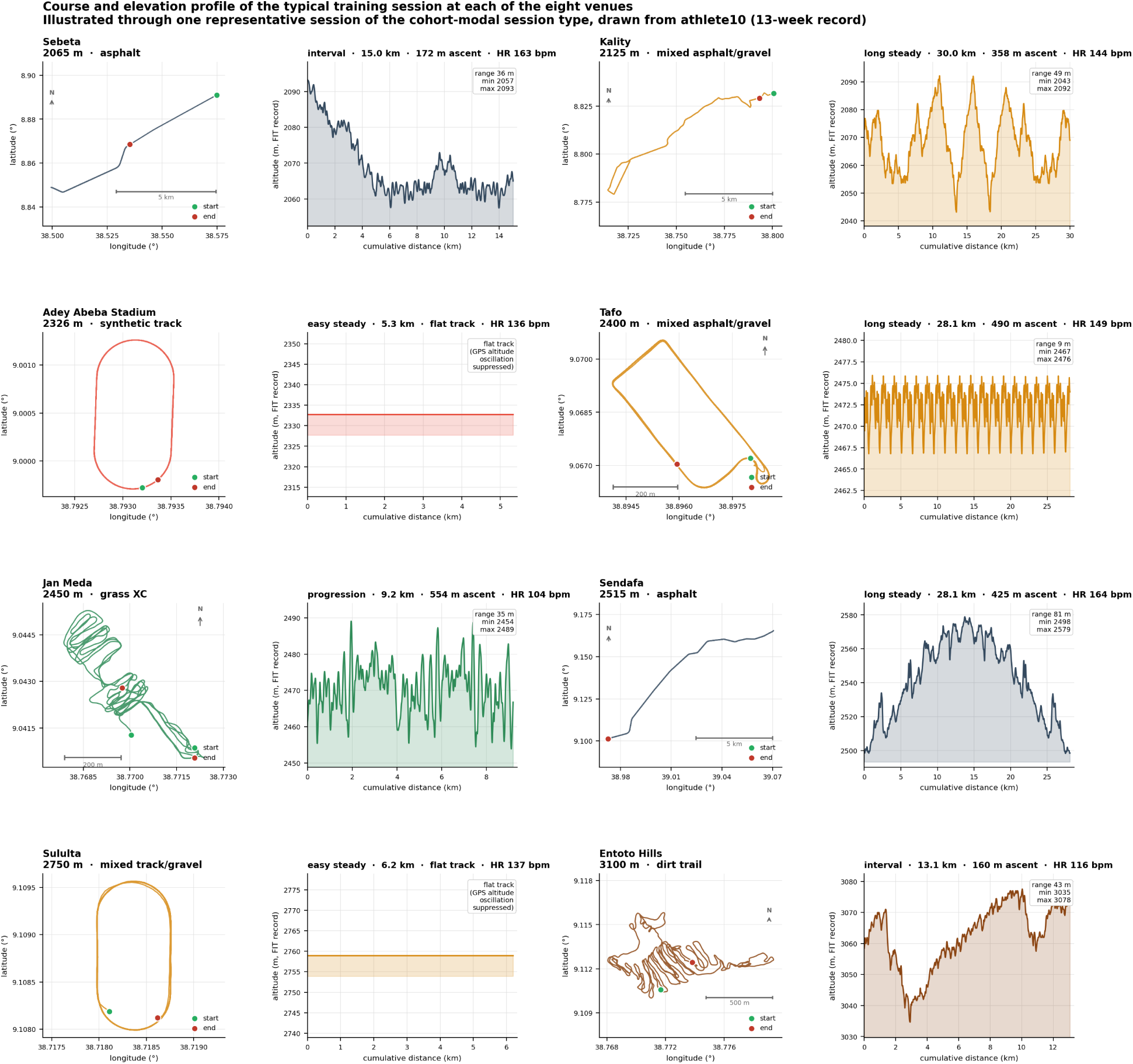
A representative course and elevation profile at each venue. The tracks are flat, the long-run roads gently rolling, and the high trails continuously climbing or undulating. On the climbing trails a steady effort rises and falls with the ground, so an automatic classifier labels it as interval work even when it is one continuous push.

### Training volume and weekly structure

The cohort runs a median of about 119 kilometres a week, ranging from 64 for the 10 km specialist to 161 for the highest-volume marathoners, across roughly eight and a half sessions (Figure 7). Bout frequency ranges from 5.0 to 10.5 a week across athletes. The recording window falls in the competitive season, and the median barely changes when the lighter taper and recovery weeks are removed.

**Figure 7.**
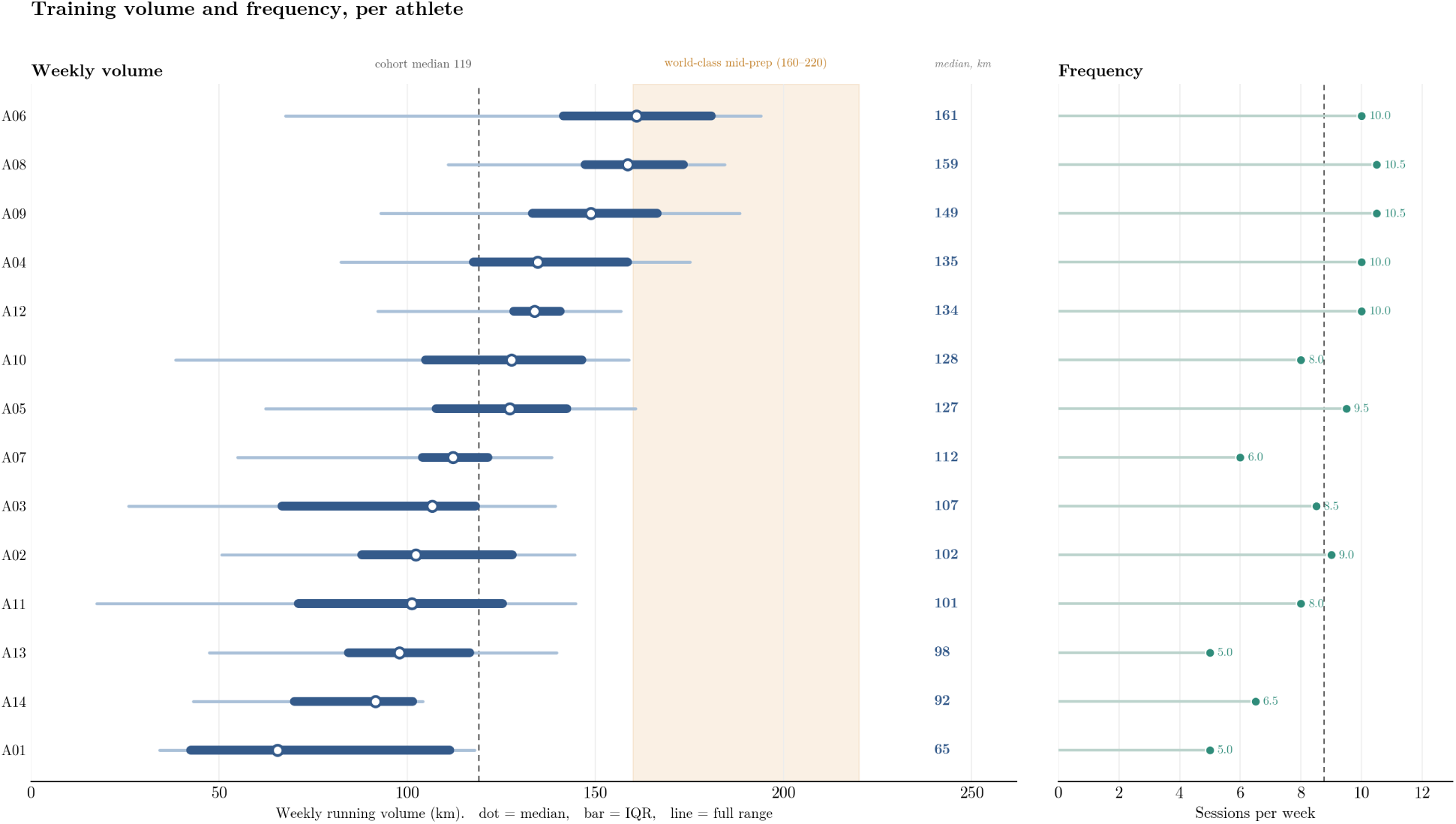
Weekly running volume and session frequency per athlete, ordered by median weekly volume. For each athlete the dot, bar, and line give the median, interquartile range, and full range; the shaded band marks the world-class mid-preparation range and the dashed line the cohort median. The cohort median of 119 km sits well below the world-class band, with only the highest-volume athletes approaching it, consistent with a competitive-season recording window.

About forty percent of training days include two runs, the rest single sessions. On the double days the morning session is the main work, around sixteen kilometres, and the afternoon is a short recovery jog of roughly half that distance at a markedly lower heart rate. The week has a steady rhythm (Figure 8). Sunday is almost always rest, in about eighty percent of athlete-weeks, and Tuesday, Thursday, and Saturday carry the heaviest work, a median of 21 to 22 km against 13 to 15 km on the lighter Monday, Wednesday, and Friday. The race taper is a sharp drop in volume, with the hardest work falling away first, compressing a roughly 128 km week to about 77 km the week before a race and to about 21 km in race week.

**Figure 8.**
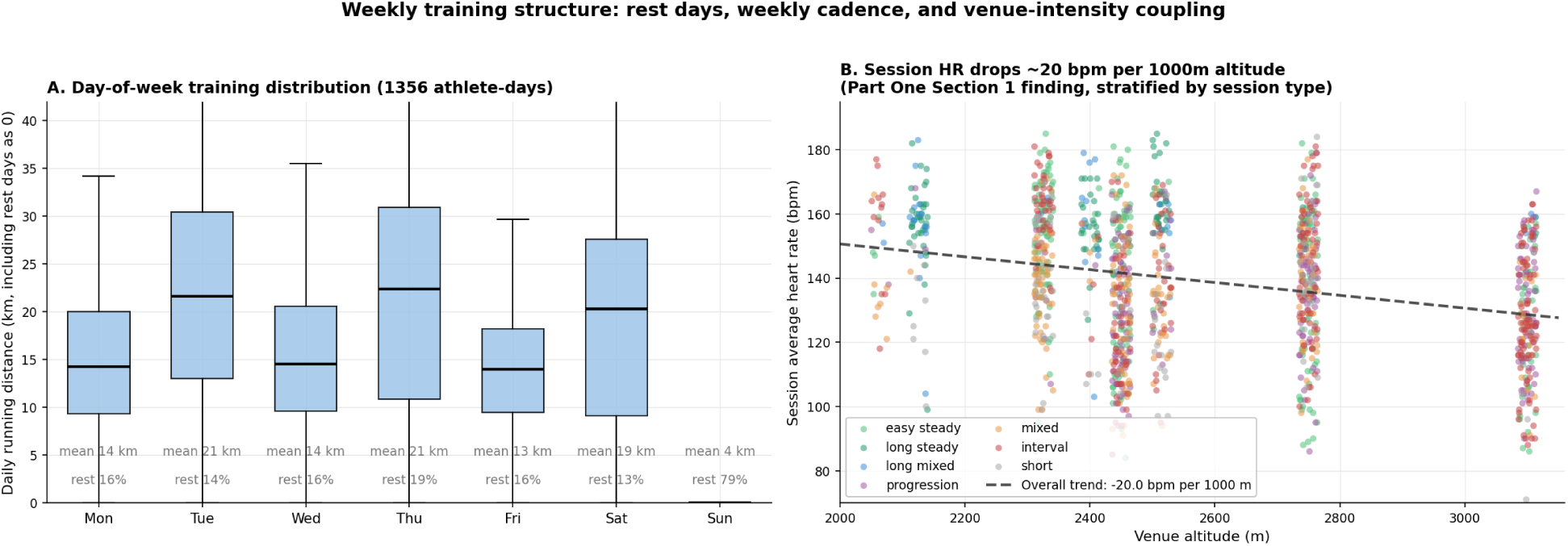
The weekly pattern: running volume by day of week and the hard-easy alternation, with Sunday rest and the heaviest work on Tuesday, Thursday, and Saturday. This rhythm, and the sharp drop in volume at the race taper, follow descriptions of world-class endurance practice.

### Intensity distribution

The share of training time spent at low, moderate, and high intensity was computed under five demarcations, four anchoring the zones to pace under different altitude corrections and one to heart rate. The distribution is relatively conventional for elite distance running, a broad base of easy running with a small moderate share and a modest amount of hard work. Once the altitude correction is applied, the easy share sits between about 75 and 93% on the pace demarcations and about 62% under the heart-rate zones, the moderate share is small at about 3 to 5% on pace and about 15% on heart rate, the hard share runs from about 4 to 23%, and the polarisation index sits above 2.0 on the three corrected pace demarcations and just below it on the heart-rate demarcation, so the corrected picture leans polarised. One demarcation stands apart. The pace scheme with no altitude correction places almost all running in the easy zone, about 98%, and leaves the moderate and hard zones nearly empty. That result is degenerate, because applying the sea-level reference paces directly to altitude-slowed running misclassifies moderate and hard work as easy, and it is the altitude correction that brings the pace-based picture back into line with the heart-rate picture. Pace and heart rate then agree closely, except on the moderate share, which the heart-rate scheme places higher. Figure 9 shows the split under each demarcation.

**Figure 9.**
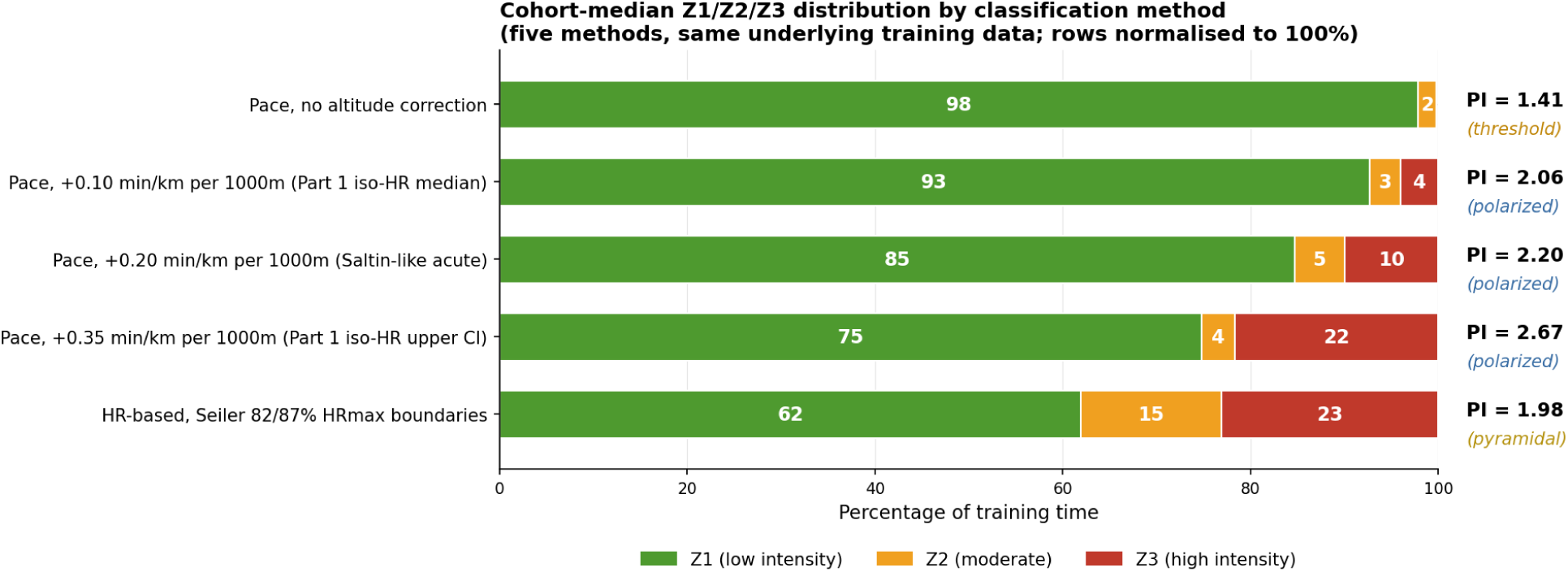
The easy / moderate / hard split under five demarcation methods, with the pace zones anchored to the reference pace scale (see Methods). Once the altitude correction is applied the distribution leans polarised, dominated by easy running with only a small moderate share; the uncorrected pace scheme is degenerate, placing almost all running in the easy zone.

### Intensity without fixed zones

The recorded paces were also binned directly, with no zone scheme applied. Each segment’s pace was adjusted to altitude and expressed relative to its athlete’s marathon pace, pooled across the fourteen runners, and summed into five-second bins (Figure 10).

**Figure 10.**
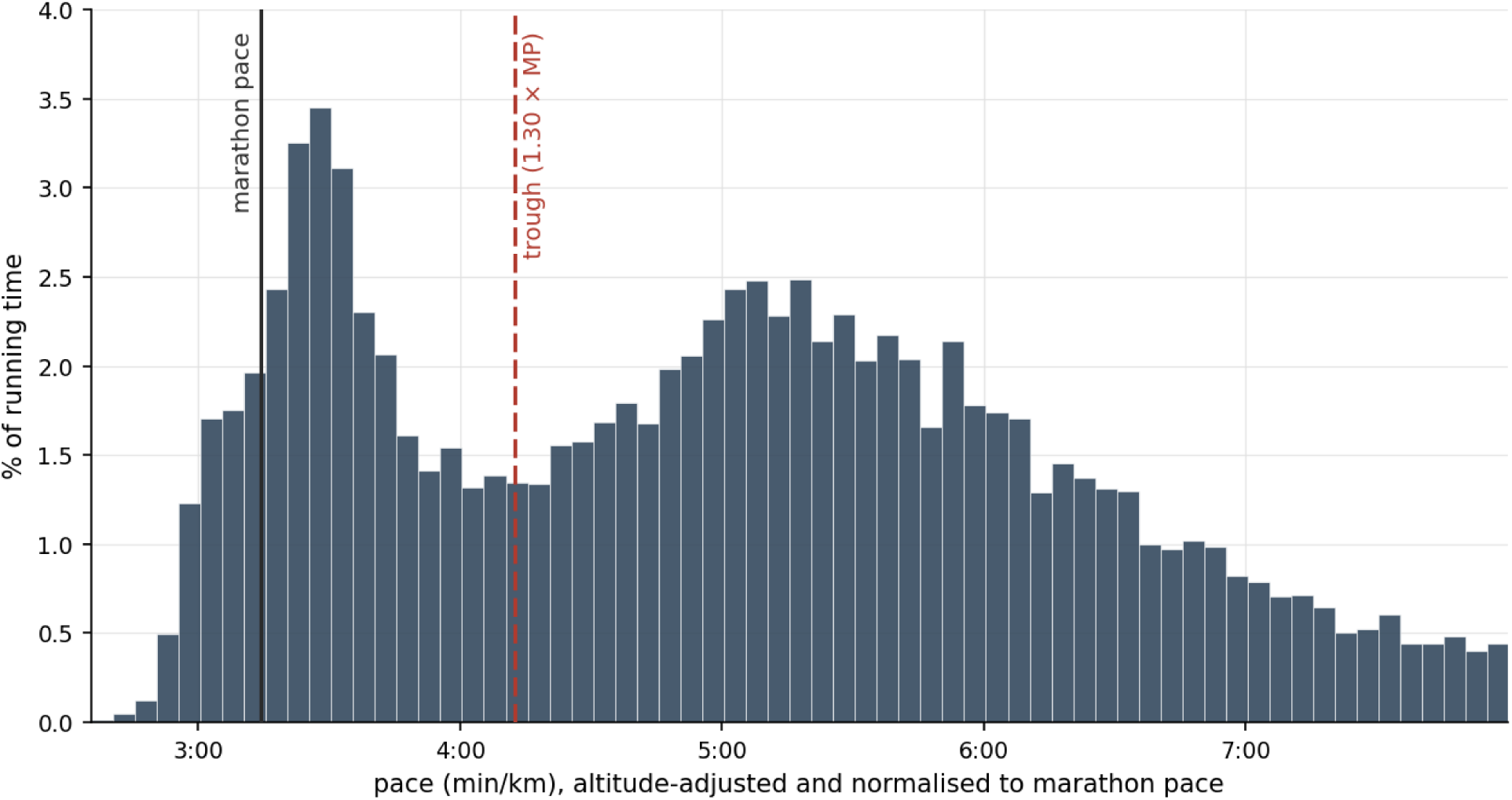
The distribution of running time by pace, with no intensity zones applied. Each segment’s pace is adjusted to altitude and normalised so that every athlete’s marathon pace aligns to the cohort median of 3:14 min/km, allowing the fourteen runners to be pooled, and the running time is summed into five-second bins. The solid line marks marathon pace and the dashed line the trough between the modes, at a pace-to-marathon-pace ratio of 1.30, which the lattice (Figure 13) also uses as an intensity boundary. The running time falls into two modes, a faster one close to marathon pace and an easy one well slower, both slower than marathon pace.

The distribution has two modes, a faster one a little slower than marathon pace and an easy one well slower than it, with a sparse trough between them, and both modes lie slower than marathon pace. The two modes are present when interval sessions are excluded, and eleven of the fourteen athletes show the same dip individually. The faster mode is run at a higher average heart rate than the easy mode.

### Group dynamics

About three quarters of all sessions are run together, typically around nine athletes in a group. The long endurance runs and the harder sessions are almost always group sessions. Only the progression-style sessions, structured solo efforts that build pace through the run, are commonly done alone. The group fraction is highest for the long and hard sessions, about 93% of long steady runs, 97% of long mixed runs, and 80% of intervals, and lowest for easy and progression work, near 55% and run in smaller groups.

Within the long group runs the athletes settle into two pace bands separated by about half a minute per kilometre (Figure 11), and the faster athletes are reliably in the faster band, the placement tracking each athlete’s fitness closely (Figure 12). The structure comes from 12 tightly synchronised long group runs of five or more athletes, where splitting into the two subgroups cuts the within-run pace spread by about 62%, to a tight 0.08 min/km within a band. The fitness link is quantified by a Spearman correlation of −0.62 between a runner’s share of time in the faster band and their gap to the world record (p = 0.017, n = 14, bootstrap interval −0.86 to −0.19).

**Figure 11.**
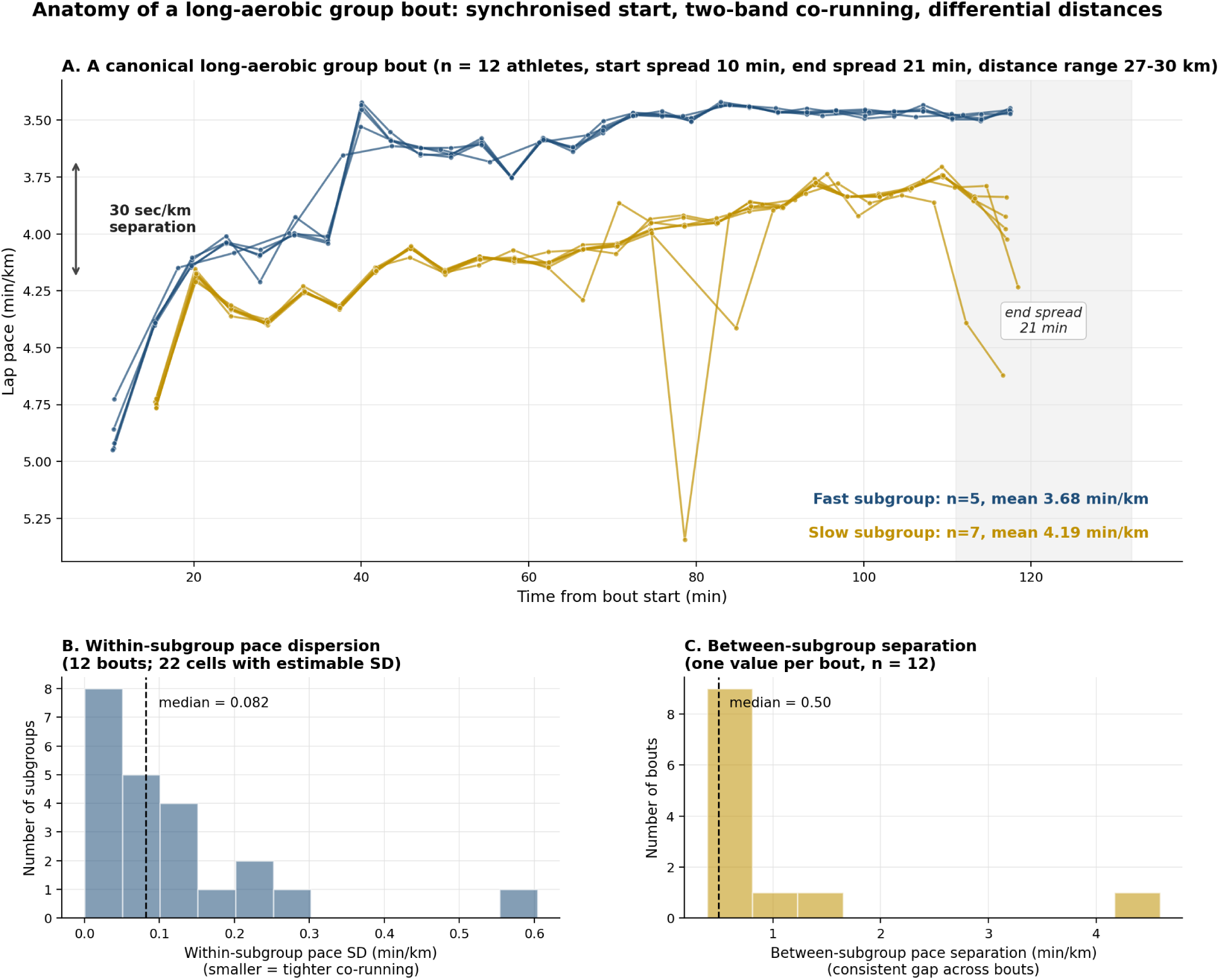
Within a single long group run, the athletes separate into a faster and a slower pace band, here about half a minute per kilometre apart, while running the session together. A single group run therefore delivers a different dose to each runner without an individual plan.

**Figure 12.**
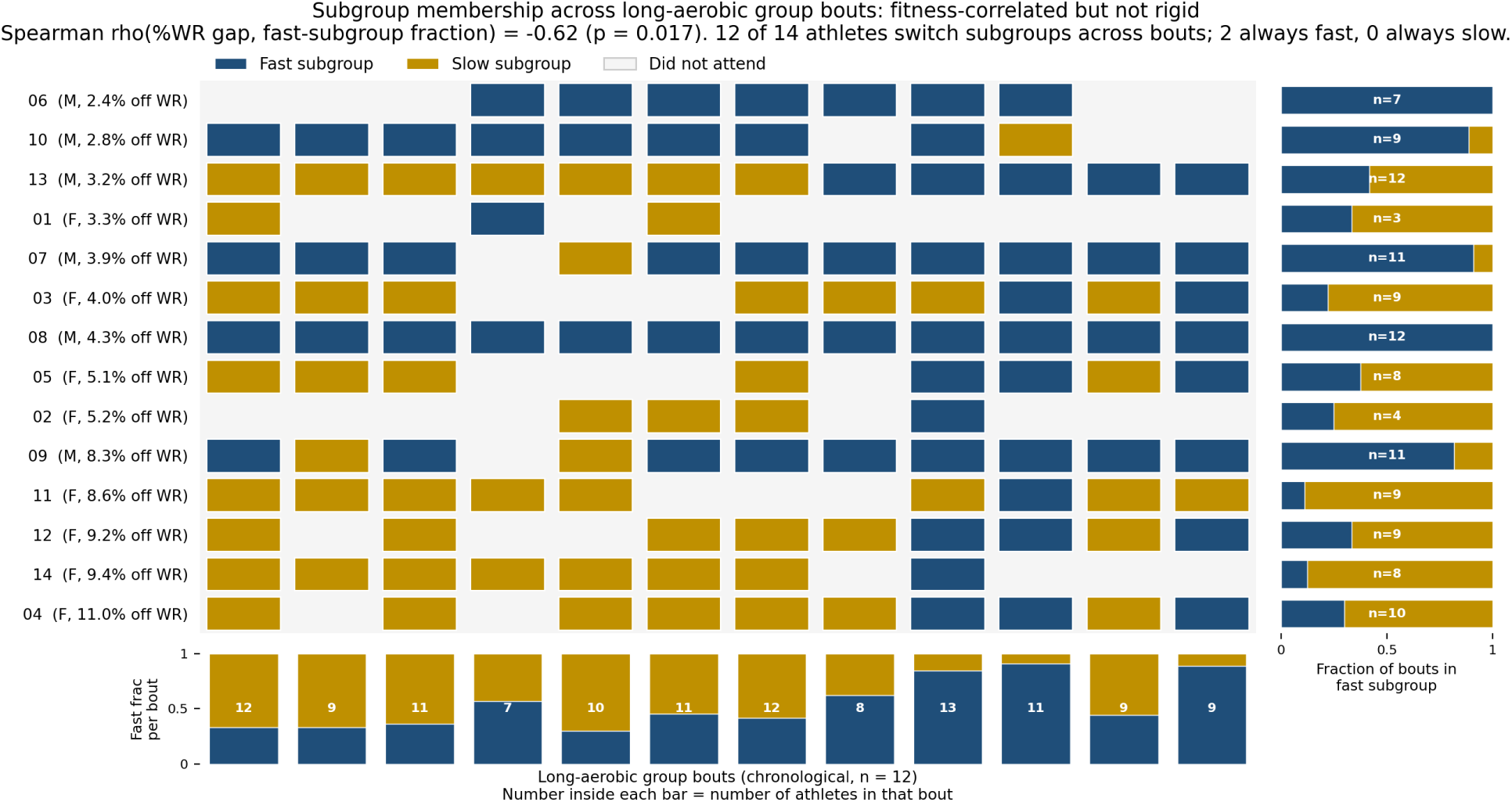
Across long group runs, band membership tracks fitness: the faster athletes, those with the smaller gap to the world record, are reliably in the faster band.

### The training as a multidimensional map

The training is described here as a five-dimensional object, with axes of altitude, surface, intensity, session structure, and session duration, each session sitting at one position on all five. Because five axes cannot be drawn together, the object is rendered as three lattices that share a grid of altitude band (low, medium, or high) and surface (soft, track, or road), each adding one of the remaining three axes. In every lattice the altitude-and-surface grid is 3×3 and the added axis is split into three panels, a 3×3×3 layout of twenty-seven combinations of which twelve have no venue in this cohort’s catchment. The three are read together as one compressed map of the training.

The first lattice shows intensity, set by effort, dividing the running at marathon pace and at the trough of the zone-free distribution (Figure 10) into an easy base slower than the trough, steady endurance running between the trough and marathon pace, and quality work faster than marathon pace. Three altitude-and-surface combinations hold about 84% of all training time, a share that does not depend on how the effort axis is drawn (Figure 13). By effort the easy base is about 65% of training time, the steady work about 27%, and the quality work about 8%. The base is mostly the high-altitude soft-trail running, 36% of all training time at about 71% of maximum heart rate and a pace near 5:40 min/km. The steady work is on the roads, 10% of training time on the mid-altitude roads and 7% on the low-altitude roads. The quality work is on the mid-altitude track and road.

**Figure 13.**
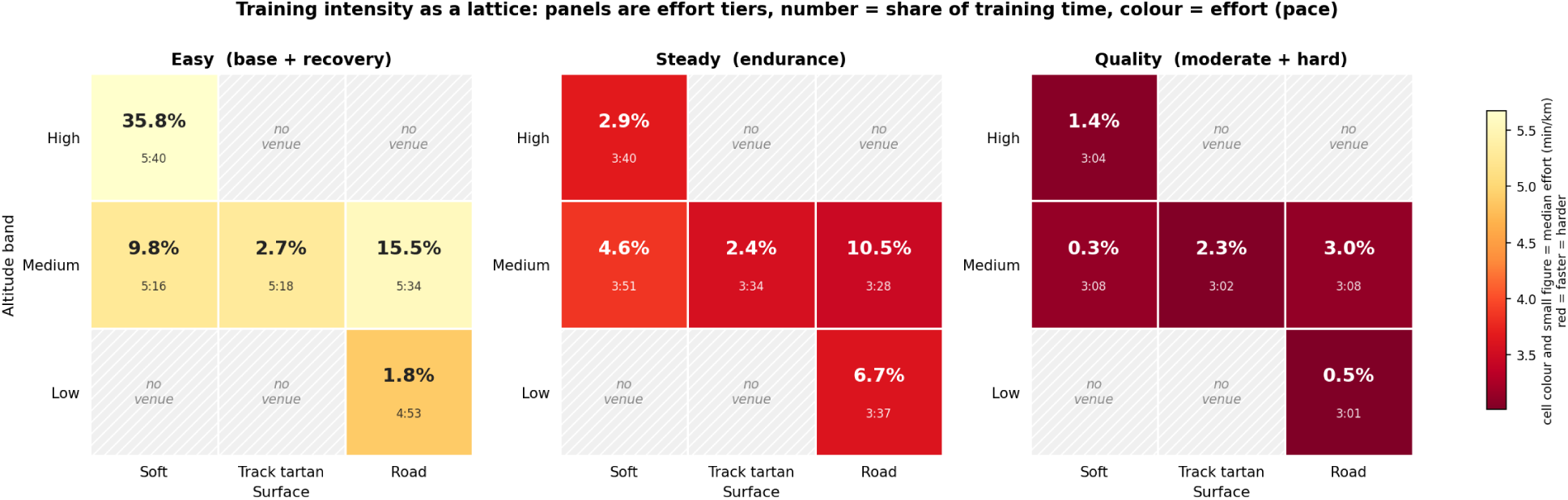
Training as a 3×3×3 lattice of intensity, altitude band, and surface. The intensity tier follows the two modes of the zone-free distribution (Figure 10), an easy base slower than the trough, steady endurance work between the trough and marathon pace, and quality work faster than marathon pace. In each cell the large figure is its share of total training time and the smaller figure, matched by colour, is the median altitude-corrected pace, redder being faster. Twelve of the 27 combinations have no venue nearby. The easy base holds about 65% of training time, the steady work about 27%, and the quality work about 8%.

The second lattice shows session structure, the shape of the pace within a session (Figure 14). It separates even running, smoothly varying running of the kind a climb or a progression produces, and the alternating running of interval and fartlek work. Continuous, even-paced running sits on the roads. The smoothly varying running is overwhelmingly on the high soft trails, where the rolling ground turns a steady effort into continuously rising and falling pace. The alternating rep and fartlek work is spread more widely, across the high trails, the mid-altitude roads, and the synthetic track. As whole-session running time these are about 39%, 40%, and 21% in turn. Continuous running carries the same low pace variability on soft ground as on road, so its evenness reflects how it is run and not the ground beneath it.

**Figure 14.**
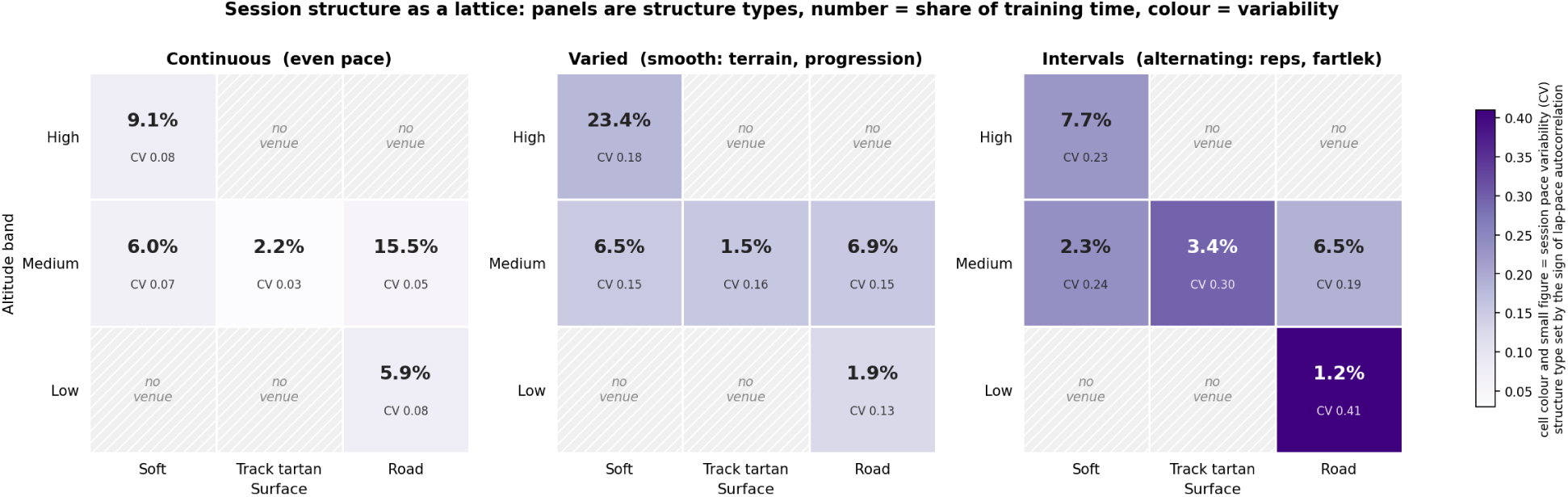
The same sessions as a lattice of session structure, altitude band, and surface. Sessions are grouped by how much their pace varies and in what pattern into continuous even running, varied running that trends smoothly under terrain or a progression, and interval-like rep and fartlek running that alternates fast and slow. In each cell the large figure is its share of total training time and the smaller figure, matched by colour, is the median session pace variability. Because the laps are one-kilometre automatic splits, sub-kilometre terrain averages out, so continuous running reads as low variability on all surfaces.

The third lattice shows session duration (Figure 15). The shortest sessions, a median near 22 minutes, are on the track, and the longest, near 90 minutes, on the low-altitude roads, with the high soft ground near 65 minutes and the mid soft and road venues near 50 minutes. As whole-session time about 18% of training is in short sessions, 36% in medium, and 45% in long. Read together, the three lattices place intensity, structure, and duration on the same altitude-and-surface grid, so each venue has a full profile of how hard, how varied, and how long its running is. The same three lattices can be resolved in time rather than pooled over the window (Figure 16).

**Figure 15.**
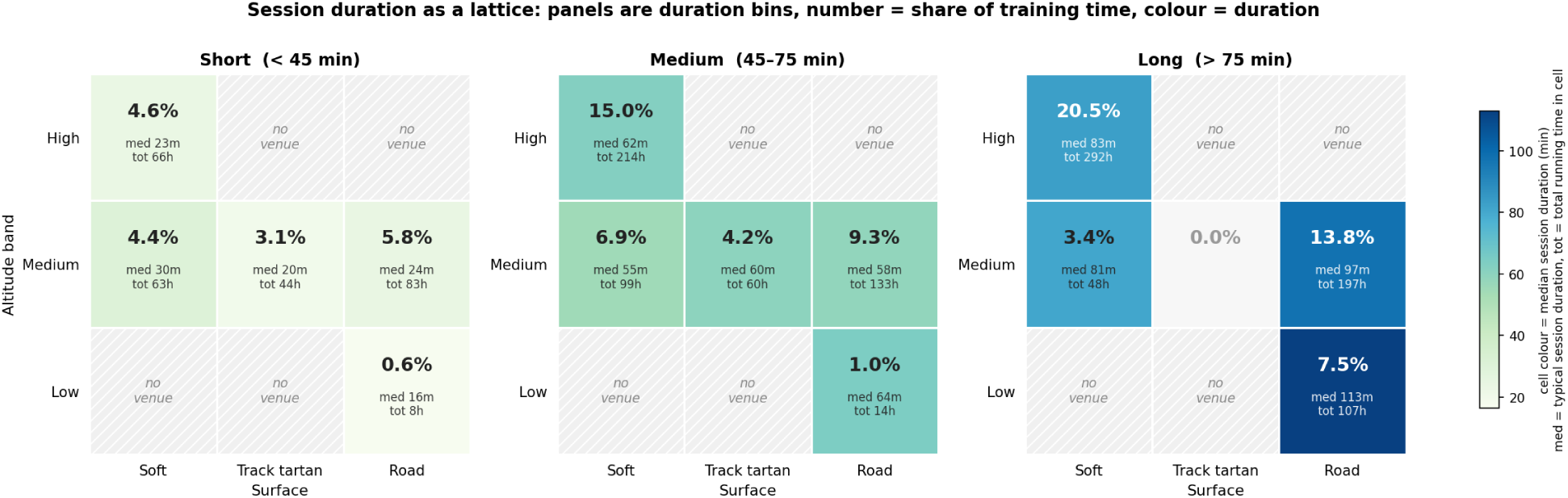
The same sessions as a lattice of session-duration bin, altitude band, and surface. Sessions are grouped by duration into short under 45 minutes, medium from 45 to 75, and long over 75. In each cell the large figure is its share of total training time; beneath it, med is the median session duration and tot the total running time accumulated in that cell, and the colour follows the median session duration, darker being longer.

**Figure 16.**
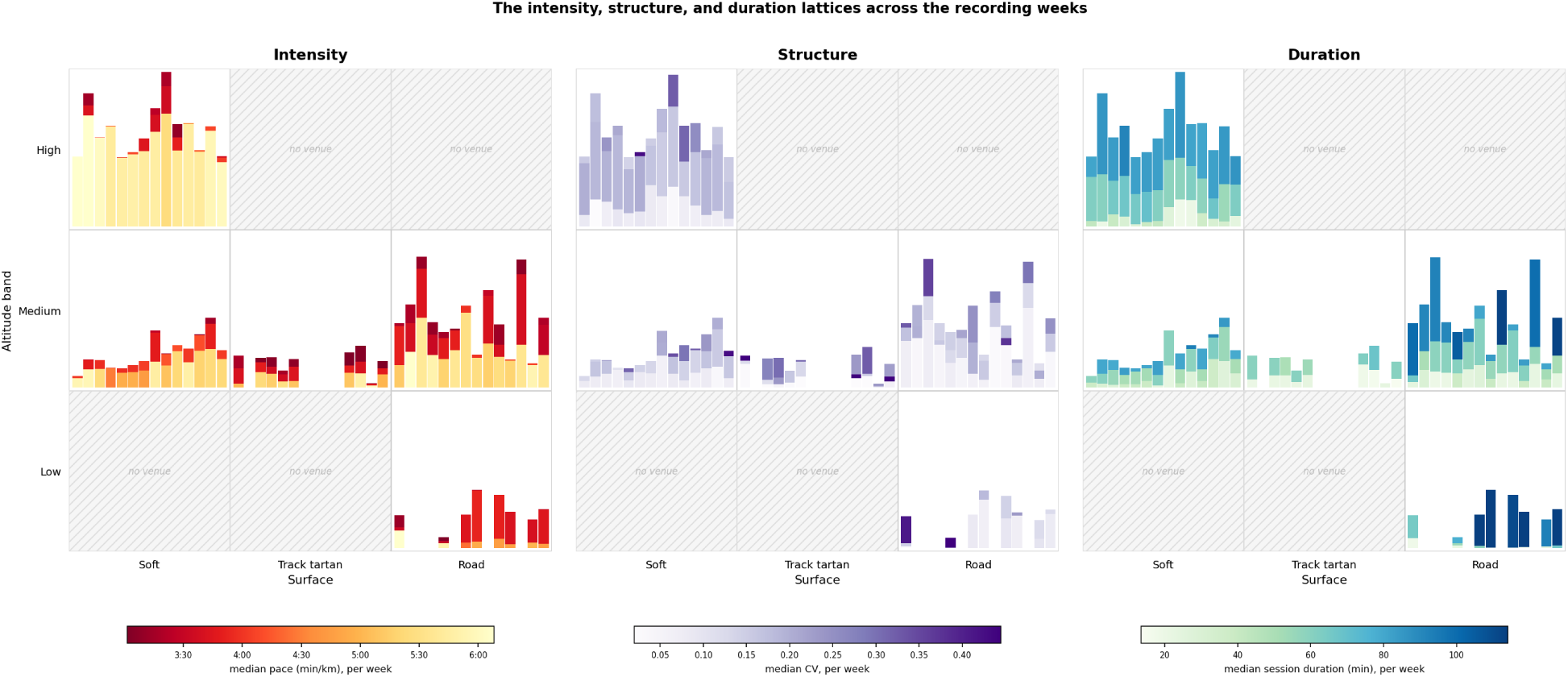
The intensity, structure, and duration lattices of Figures 13 to 15 unfolded across the fourteen recording weeks. Each cell of the altitude-and-surface grid holds a weekly stacked bar rather than a single value. The bars run left to right by week, the stacked segments are the cell’s bins by running time (easy, steady, and quality for intensity; continuous, varied, and interval for structure; short, medium, and long for duration), the bar height is running time per athlete active in that week on a scale shared by every cell so that heights are comparable, and each segment’s colour is that week’s median pace, variability, or session duration. Summing a cell over the weeks reproduces its value in Figures 13 to 15, and the same decomposition can be taken at any temporal resolution and for a single athlete.

Figure 16 unfolds the three lattices across the fourteen recording weeks, each cell carrying a weekly stacked bar in place of a single value. The running time in a cell is not steady across the weeks, varying in both amount and composition. The easy base on the high soft trails is present in every week and is the largest and most regular contribution, while the track work and the long runs on the low-altitude roads appear in some weeks and not in others. Collapsing each cell over the weeks returns the values of Figures 13 to 15.

## Discussion

This study describes the training of one cohort of elite Ethiopian distance runners and, secondarily, examines how an AI-assisted workflow contributed to that description. The principal empirical finding is that the cohort’s training is organised along several dimensions at once. Session purpose, effort, altitude, surface, terrain, and group execution are combined, and a single axis of intensity does not capture the structure. Long aerobic running, trail running, structured repetitions, and recovery work occur in different places, and those places each carry their own combination of altitude, ground, and terrain. The result is a training system in which intensity is embedded in venue rather than prescribed independently of it.

This structure helps explain why the initial pace analyses required caution. Pace could not be compared directly across venues, because the same effort produces different speeds at different altitudes and on different surfaces. The matched-heart-rate altitude estimate, approximately +0.10 min/km per 1,000 m, is conservative relative to the wider range of estimates produced by less controlled comparisons and is consistent with the expectation that long-term altitude-adapted athletes show a smaller decrement than non-adapted runners [2]. Surface introduced a separate difficulty. In this dataset, surface, altitude, terrain, and venue are not independently distributed. Each surface belongs to a particular place, so a pooled model cannot cleanly isolate a “surface effect” from the broader conditions of that venue. This is a feature of the coaching structure as well as a statistical limitation. The athletes do not select altitude, surface, and terrain independently. They train at venues that combine them.

The route-overlap and terrain analyses sharpen this interpretation. Tracks and small loops are highly repeatable, road sessions follow broadly similar courses at different lengths, and trail sessions vary much more from one outing to the next. Each venue’s terrain is itself part of the training. The flat track allows exact structured repetition, the gently rolling roads allow a continuous aerobic effort to be held over long distances, and the climbing trails turn a steady effort into continuously varied work that rises and falls with the ground [3]. The ground is therefore a training variable in its own right, and a venue label or an average pace cannot recover what it contributes. Interpreting the sessions correctly requires the kind of ground covered, the terrain profile, and the way effort changes with the landscape.

The cohort’s weekly volume is lower than the published figures for world-class distance running, but the comparison is not like for like. The median of about 119 km sits below the 160 to 220 km reported for world-class runners in mid-preparation [6], and the recording window falls in the competitive season, when volume drops from its preparation peak [7]. The median reflects an ordinary competitive-season load rather than a taper artefact. Other features match the literature closely. The weekly rhythm, with the heaviest work on Tuesday, Thursday, and Saturday and Sunday almost always rest, is the hard-day, easy-day alternation reported as near-universal in elite endurance training [6], and the race taper, a sharp drop in volume with the hardest work falling away first, follows published descriptions of world-class practice.

The intensity results are informative and they look relatively conventional. On the half-marathon and marathon pace scale used for world-class distance runners [6], with the data-derived altitude correction applied, the distribution is dominated by easy running, has only a small moderate share, and leans polarised, falling in the same region as published altitude training logs, including the widely reported logs of Eliud Kipchoge, in which easy running accounts for roughly 82 to 91% of the work, moderate for about 3 to 10%, and hard for about 6 to 8% [6]. The cohort’s polarisation index spans 1.41 to 2.67 across the five demarcations, with the three corrected pace demarcations above 2.0, placing this cohort with the polarised distributions reported for world-class runners and above the pyramidal and threshold distributions reported for subelite groups (Figure 17). The heart-rate classification gives the same broad picture, although it places somewhat more running in the moderate zone, a divergence between race-pace and physiological zoning also reported elsewhere [15].

**Figure 17.**
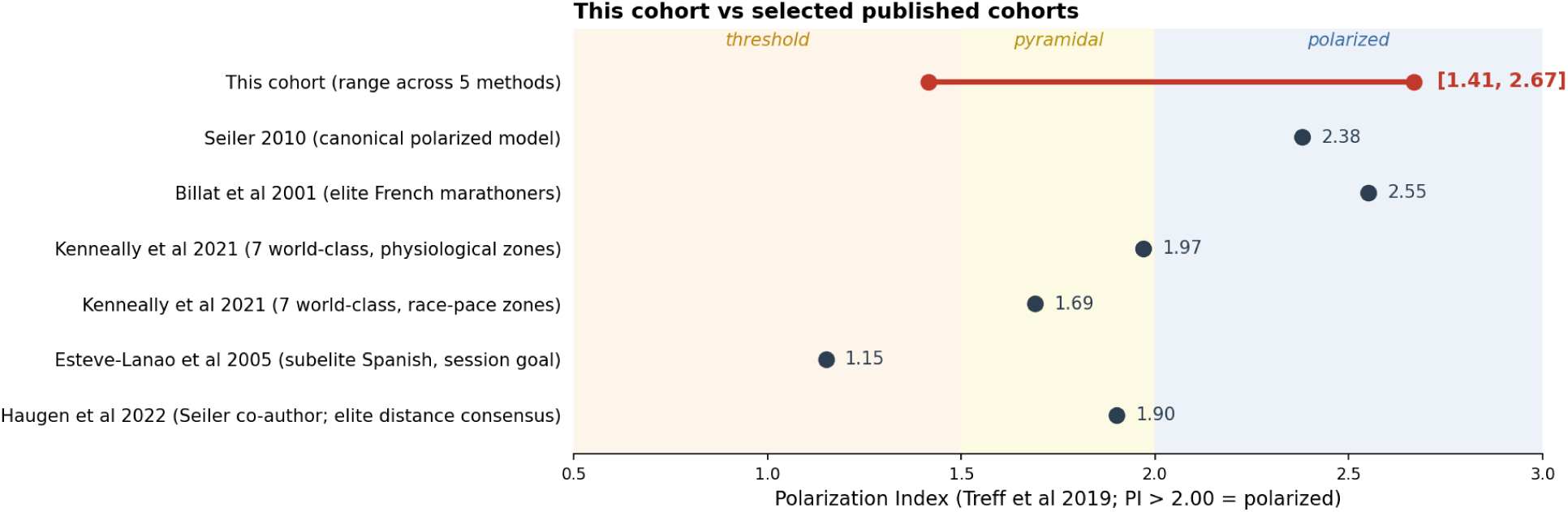
The cohort’s polarisation-index range across the five demarcation methods, set against selected published cohorts. The corrected pace demarcations place this cohort with the polarised distributions reported for world-class distance runners; the heart-rate demarcation sits just below the polarised boundary.

The cohort does not look unusual. A training-intensity distribution is a one-dimensional projection of a multidimensional practice though. It shows how much training falls into each zone, but not where that work is done, on what surface, over what terrain, or how the venue changes its meaning. Altitude and surface modify the meaning of pace, and terrain modifies the meaning of effort, so a single intensity-distribution label gives an incomplete account of the practice. The zone-free analysis points in the same direction. When corrected paces are binned directly, without imposing intensity thresholds, the running separates into two modes, one clearly easy and one faster and closer to marathon pace, with a sparse middle between them. The faster mode is also run at higher heart rate, so the modes differ in effort as well as speed. The small moderate share therefore reflects a trough between preferred pace-effort regions, while the lattice shows where those regions sit within the altitude-surface structure of training, with the harder work placed differently from the easy bulk.

The group-session analyses add a further dimension. In many European and North American models of elite endurance training, individualisation is typically described through athlete-specific zones, pace targets, laboratory testing, and written prescriptions [6, 16]. The watch data from this cohort show little evidence of that kind of individual prescription. Instead, most key sessions are run together [3]. Within long group runs, the athletes separate into two pace bands that are consistently related to performance level. A single group session can therefore deliver different training doses to different athletes without the data showing separate individual plans. The mechanism cannot be fully resolved from watch data alone. The bands may be set by the coach, negotiated by the athletes, or emerge during the run. The regularity is nonetheless clear. Group execution produces a structured form of individualisation.

The multidimensional framework brings these findings together. Classifying sessions at once by altitude, surface, intensity, session structure, and session duration makes visible features that no single axis can show. The entanglement of altitude and surface arises because each venue fixes both. The conventional-looking intensity distribution turns out to be a projection that hides where the work is placed, because a given pace means different things at different altitudes and on different surfaces. The concentration of training time into a small subset of the available combinations suggests that the venues are chosen deliberately rather than used opportunistically, and even the small share of moderate and hard work is placed on different ground from the easy bulk. Easy, moderate, and hard work are distributed through particular combinations of altitude and surface available in this environment. Session structure and duration sort by venue in the same way. The continuous steady running is on the roads, the smoothly varying running on the high trails, and the alternating rep and fartlek work spread across the trails, roads, and track, so much of the easy base is built through the rolling trail running, and the longest sessions fall on the roads and high trails while the track work is short. This quantitative account gives empirical, GPS-based form to a characterisation of East African training that has so far been made only in conceptual terms, as a variability-driven and terrain-embedded practice rather than one organised along a single controlled-intensity axis [3].

A lattice value is an average over the whole window, but the decomposition behind it is not fixed at that scale. Every cell is built from dated, athlete-labelled sessions, so the same map can be unfolded to any resolution, from a single day through a week, month, or season to a full training cycle, and read for the cohort or for one athlete (Figure 16). Seen this way the training is not stationary across the block, the volume and composition within a cell moving from week to week, so the lattices describe how the training is arranged at a given time as much as on average. The framework therefore reaches periodisation and individual monitoring without any change to its five axes.

This interpretation should not be taken as evidence that the coaches explicitly use a formal multidimensional model. The lattices are an analytic description of the structure recovered from the data, not necessarily a representation of the coaches’ own language or conscious planning system. The coaches’ practice is more likely to be experiential, contextual, and responsive to the athletes and terrain. The value of the framework is that it translates part of that practice into measurable form. It shows how a coaching tradition based largely on experience can leave a stable footprint in GPS-watch data, even when the reasoning that produced the pattern remains outside the dataset.

The workflow used in this study also has methodological implications, and the division of labour follows the vocabulary set out in the Methods. The AI tools were useful for searching the data, generating candidate analyses, writing and checking code, drafting text, and identifying errors during independent review. They helped make a detailed dissection of a large and complex training record feasible. This is work at the associative rung of Pearl’s framework [8], finding and testing patterns in what was recorded. Directing the analysis toward a particular question or place, such as whether repeat sessions share ground or how the group sessions are run, is intervention, the middle rung, where the investigators set the direction and the AI carried it out once pointed. Supplying the organising framework, the decision to view the training as a multidimensional object rather than along a single intensity axis, sits just above the top rung, and is the one move a model does not make on its own, because it recombines what its training already contains and does not reach for terms from outside it [9]. The autonomous searches and the guided analyses surfaced and tested the regularities; that framework came from an investigator familiar with elite endurance training. On this evidence, AI accelerated and broadened the empirical work, while direction, interpretation, and the final claims remained the investigators’ responsibility.

The study therefore contributes a detailed description of how this cohort trained, not a causal explanation of why Ethiopian runners succeed. Much of what was observed is consistent with prior accounts of elite East African and world-class distance running, which describe high but seasonally moderated volume, hard-easy weekly alternation, substantial easy running, altitude exposure, and group training. The present data add a more precise account of how those elements are combined in practice. In this cohort, training is shaped by the pairing of intensity with altitude and surface, by the use of terrain as part of session design, and by group execution as a mechanism of individualisation. These features are difficult to see through models that reduce training primarily to weekly volume and intensity distribution.

The broader implication is that high-resolution training data can recover the measurable structure of a coaching practice that has not been formally written down. It cannot, by itself, recover the tacit judgement behind that practice, such as why a coach chooses one venue on a given day, how an athlete’s readiness is assessed, or how a session is adjusted in response to weather, terrain, group dynamics, or race timing. The framework described here should therefore be treated as a descriptive account of this cohort in this setting.

Its value lies in showing what standard categories miss when they are applied without sufficient attention to place, surface, terrain, and group organisation, rather than in turning the practice into a portable formula.

### Limitations

This is one cohort of elite Ethiopian distance runners observed across a single competitive season, without a comparison group or laboratory measurement, so the findings describe how this group trained and cannot be used to establish causality or to generalise directly to other athletes. Training-watch data records what was done but not why, and cannot see the coach’s instruction, the athlete’s intent, or the reasoning behind a session, so the prescription system is inferred from its footprint rather than observed at its source. The lattice is a descriptive framework, and its intensity tiers depend on the altitude pace-correction, which is itself uncertain; the corrected demarcations agree that the training is easy-dominated and leans polarised, but the exact split between the tiers shifts with the correction, so the tier boundaries should be treated as approximate. The same correction sets the effort shown by cell colour in Figure 13, so that colour shifts with it in absolute terms, though the ordering of cells by effort held across the corrections tested. The variability and duration lattices are computed over whole sessions, so their venue shares omit the small fraction of running in very short or venueless sessions, and the one-kilometre lap resolution understates the pace variability of short track repetitions. The contrasts with Western and Scandinavian practice rest on the published literature, since the study has no matched comparison cohort of its own. Finally, the structure recovered here is rooted in this group’s circumstances, the altitude they live at, the venues open to them, and the way they train together, so it describes a particular practice in a particular place.

### Conclusions

This study reconstructs how one group of elite Ethiopian runners trained from the data recorded by their GPS watches. The central finding is a practice organised simultaneously across five dimensions of altitude, surface, intensity, session structure, and session duration. This structure was executed largely by effort, shaped by the available training venues, and mediated through the group. The intensity distribution reproduces the conventional picture of elite distance running, yet a single-axis summary captures only part of this practice and misses how hard, moderate, and easy work are paired with particular places, surfaces, and terrains. The coaches responsible for this cohort have long worked with runners at the elite and world-class level, and their system appears to have developed through years of practical experience in a physical and cultural setting that does not map neatly onto the codified structures commonly used in endurance-training science.

The centre of the study is the passage from tacit knowledge to measurable pattern. The coaches’ tacit knowledge, formed through experience with these athletes, venues, surfaces, altitudes, and group dynamics, shaped the training prescription. That prescription shaped the athletes’ day-to-day practice, and the practice left a footprint in the GPS-watch data. The AI-assisted workflow helped recover that footprint as a set of regularities. They included where the athletes trained, how effort was paired with altitude and surface, how terrain affected the interpretation of sessions, and how group running shaped individual training dose. The investigator then supplied the framework that made those regularities intelligible as one organising structure, in which altitude, surface, intensity, session structure, and session duration are considered together.

The lattices therefore record the visible structure of the training, not the full coaching knowledge that produced it. They capture what the data can measure, while leaving out many of the judgements that matter in practice. These include how the coach reads an athlete on a given day, why one venue is selected over another, how the group is managed, and how the prescription changes with weather, fatigue, race timing, and individual readiness. The framework can be written down, checked, and shared as a description of this cohort’s training structure. Any use beyond this cohort would need to be grounded in the tacit knowledge of the new setting, including its athletes, coaches, venues, and constraints.

The same sequence gives the paper its point about AI-assisted research. AI and quantitative analysis can expose patterns, test claims, and make parts of the research process reproducible. In this study, however, their value depended on human domain expertise at both ends, the coaches whose experience shaped the training and the investigator whose experience made the recovered pattern interpretable. Rather than marginalising that expertise, better instruments made its role easier to see.

## List of abbreviations

- **AI:** Artificial Intelligence (used throughout to mean generative AI, i.e. large language models and the tools built around them)
- **ERA5:** Fifth-generation European Centre for Medium-Range Weather Forecasts atmospheric reanalysis
- **FIT:** Flexible and Interoperable Data Transfer (GPS-watch activity file format)
- **GPS:** Global Positioning System
- **GPT:** Generative Pre-trained Transformer
- **STROBE:** STrengthening the Reporting of OBservational studies in Epidemiology

## Declarations

### Ethics approval and consent to participate

Ethical approval was obtained from the IRB of the College of Natural and Computational Science, Addis Ababa University Ref No: EDRE/04/2017/2025.

### Consent for publication

Not applicable. All data are presented anonymously.

### Availability of data and materials

Raw FIT files, cleaned segment-level dataset, ERA5 dataset, data dictionary, verification scripts and citations audit supporting this manuscript are available from the corresponding author upon reasonable request, subject to athlete privacy protections.

### Supplementary information

Supplementary Information accompanies this paper. It reports the resource cost of the human-AI workflow, including investigator time, model usage, wall-clock time, and metered cost.

### Competing interests

The authors declare no competing interests.

### Funding

No funding was received for conducting this study.

### Authors’ contributions

Stephen Seiler, Abay Yisamaw, Palo Galko and Thomas Haugen conceived the study. Abay Yisamaw collected or curated the field metadata. Abay Yisamaw and Palo Galko processed the GPS data. Palo Galko designed the human-AI workflow. Palo Galko conducted the statistical analyses and verification scripting. Stephen Seiler, Abay Yisamaw, Palo Galko and Thomas Haugen contributed domain interpretation. Palo Galko, Stephen Seiler, Abay Yisamaw and Thomas Haugen contributed to adversarial review, citation verification, and manuscript revision. All authors read and approved the final manuscript.

## Data Availability

All data produced in the present study are available upon reasonable request to the authors

## Acknowledgements

The authors gratefully acknowledge the participating athletes and coaching staff for their collaboration and commitment to this investigation.

## Authors’ information

Not applicable.

## Use of generative AI and AI-assisted tools

Generative AI and AI-assisted tools were used as part of the study workflow described in the manuscript. Delv-e was used for autonomous hypothesis exploration. Claude Opus 4.7 was used under direct human supervision for claim construction, code drafting, sensitivity-check design, prose drafting, and manuscript development. OpenAI GPT-5.4 was used in an independent adversarial role to stress-test methods, statistics, prose, figures, and citations. All numerical claims retained in the manuscript were checked against verification scripts or supporting analyses. AI-generated citations were not accepted without independent verification. The authors take responsibility for the final content of the manuscript.

## Notes

### Competing Interest Statement

The authors have declared no competing interest.

### Summary of Updates

This manuscript has been revised to improve readability and clarity. The text has been condensed and streamlined to focus more sharply on the core findings and methodology.

## References

1. Wilber RL, Pitsiladis YP. Kenyan and Ethiopian distance runners: what makes them so good? Int J Sports Physiol Perform. 2012;7(2):92–102. doi:10.1123/ijspp.7.2.92.

2. Saltin B, Larsen H, Terrados N, Bangsbo J, Bak T, Kim CK, et al. Aerobic exercise capacity at sea level and at altitude in Kenyan boys, junior and senior runners compared with Scandinavian runners. Scand J Med Sci Sports. 1995;5(4):209–221. doi:10.1111/j.1600-0838.1995.tb00037.x.

3. Grivas GV, Alkasasbeh WJ, Trigonis I, Karakatsanis K, Amawi AT. Training variability and threshold density: a conceptual comparison of East African and Norwegian endurance training systems. Front Physiol. 2026;17:1878492. doi:10.3389/fphys.2026.1878492.

4. Mooses M, Mooses K, Haile DW, Durussel J, Kaasik P, Pitsiladis YP. Dissociation between running economy and running performance in elite Kenyan distance runners. J Sports Sci. 2015;33(2):136–144. doi:10.1080/02640414.2014.926384.

5. Bayissa M, Haile D, Afework D, Jebessa G, Mooses K, Mooses M. Association between running economy and VO_2_max values in high-level Ethiopian male and female distance runners measured at high altitude. BMC Res Notes. 2025;18:319. doi:10.1186/s13104-025-07397-8.

6. Haugen T, Sandbakk Ø, Seiler S, Tønnessen E. The training characteristics of world-class distance runners: an integration of scientific literature and results-proven practice. Sports Med Open. 2022;8(1):46. doi:10.1186/s40798-022-00438-7.

7. Casado A, González-Mohíno F, González-Ravé JM, Foster C. Training periodization, methods, intensity distribution, and volume in highly trained and elite distance runners: a systematic review. Int J Sports Physiol Perform. 2022;17(6):820–833. doi:10.1123/ijspp.2021-0435.

8. Pearl J, Mackenzie D. The book of why: the new science of cause and effect. New York: Basic Books; 2018.

9. Misra V. Shannon got AI this far. Kolmogorov shows where it stops [Internet]. Medium; 2026 Mar [cited 2026 May 24]. Available from: https://medium.com/@vishalmisra/shannon-got-ai-this-far-kolmogorov-shows-where-it-stops-c81825f89ca0

10. Galko P. Delv-e: autonomous data investigation powered by LLMs [Internet]. GitHub; 2025 [cited 2026 May 21]. Available from: https://github.com/pgalko/delv-e

11. Anthropic. Claude Opus 4.7 model documentation [Internet]. 2026 [cited 2026 May 21]. Available from: https://docs.claude.com

12. OpenAI. GPT-5.4 model documentation [Internet]. 2026 [cited 2026 May 21]. Available from: https://openai.com

13. McKay AKA, Stellingwerff T, Smith ES, Martin DT, Mujika I, Goosey-Tolfrey VL, et al. Defining training and performance caliber: a participant classification framework. Int J Sports Physiol Perform. 2022;17(2):317–331. doi:10.1123/ijspp.2021-0451.

14. Esteve-Lanao J, San Juan AF, Earnest CP, Foster C, Lucia A. How do endurance runners actually train? Relationship with competition performance. Med Sci Sports Exerc. 2005;37(3):496–504. doi:10.1249/01.MSS.0000155393.78744.86.

15. Kenneally M, Casado A, Gomez-Ezeiza J, Santos-Concejero J. Training intensity distribution analysis by race pace vs. physiological approach in world-class middle- and long-distance runners. Eur J Sport Sci. 2021;21(6):819–826. doi:10.1080/17461391.2020.1773934.

16. Seiler S. What is best practice for training intensity and duration distribution in endurance athletes? Int J Sports Physiol Perform. 2010;5(3):276–291. doi:10.1123/ijspp.5.3.276.

17. Treff G, Winkert K, Sareban M, Steinacker JM, Sperlich B. The polarization-index: a simple calculation to distinguish polarized from non-polarized training intensity distributions. Front Physiol. 2019;10:707. doi:10.3389/fphys.2019.00707.

18. Riegel PS. Athletic records and human endurance. Am Sci. 1981;69(3):285–290.

19. Crawley M. Tracking selves or tracking relationships? Means of measuring time amongst Ethiopian runners. J R Anthropol Inst. 2021;27(3):653–671. doi:10.1111/1467-9655.13556.

